# IsomiR Utility in Amyotrophic Lateral Sclerosis Prognostication

**DOI:** 10.1101/2025.05.12.25325848

**Authors:** Yahel Cohen, Ilan Sinai, Iddo Magen, Yehuda Matan Danino, Joanne Wuu, Andrea Malaspina, Michael Benatar, Eran Hornstein

## Abstract

**Background:** Amyotrophic lateral sclerosis (ALS) is a fatal neurodegenerative disease characterized by progressive motor neuron loss. IsomiRs are microRNA (miRNA) isoforms that arise from alternative processing or editing events during miRNA biogenesis. While isomiRs may carry distinct biological and clinical relevance, their potential as cell-free biomarkers in neurodegeneration remains largely unexplored.

**Method:** Here we investigated the prognostic utility of plasma isomiRs in ALS, using next-generation sequencing and two orthogonal statistical approaches.

**Findings:** We profiled cell-free isomiRs in 154 ALS patients from a British cohort and identified higher levels of one isomiR, let-7g-5p.t, to be associated with longer survival. This finding was independently validated in an international ALS cohort of 200 patients. let-7g-5p.t prognostic utility was comparable to that of neurofilament light chain (NfL) or miR-181.

**Conclusion:** These results establish isomiRs as a novel class of blood-based biomarkers in ALS with potential to refine prognostication in clinical trials for neurodegenerative diseases.

**Funding:** Target ALS, Israel Science Foundation (ISF 3497/21, 424/22) and the CReATe Consortium. All additional funding can be found in the Acknowledgements section of this paper.

## Introduction

Amyotrophic lateral sclerosis (ALS) is a neurodegenerative disease of the human motor neuron system. Heterogeneity of patient progression rate^1^ challenges the detection of therapeutic effects in clinical trials^2^. Biofluid-based biomarkers have garnered increasing attention as tools to monitor progression^3,4^. These include urinary p75^5^, troponin T2^6^ and microRNAs (miRNAs)^7^. Additionally, neurofilament light chain protein (NfL)^8^ is a strong predictor of the rate of disease progression, conveying meaningful prognostic information. Accordingly, we have recently demonstrated the utility of NfL in reducing the sample size required for clinical trials in a trial-like population^9^. miRNAs are small endogenous RNAs that contribute to post-transcriptional silencing of mRNA^10^. miRNAs are critical for motor neuron viability^11^ and their biogenesis is known to be dysregulated in ALS^12–16^. Previous studies have shown that cell-free miRNAs and in particular miR-181 might have prognostic value^7,17^. miRNA biogenesis is a multistep process that starts with the transcription of a primary miRNA, which is cleaved by the nuclear microprocessor complex to yield an intermediate double-stranded hairpin precursor (pre-miRNA)^18^. After export of the pre-miRNA from nucleus to cytoplasm, it is cut by Dicer, creating a single strand 22nt guide that is loaded on to the RNA-induced silencing complex (RISC)^19^. IsomiRs display sequence variations that distinguish them from the reference sequence miRNAs (reference miRNA), including 3’ or 5’ sequence changes, or sequence polymorphism^20–22^. IsomiR production is often the result of aberrant miRNA biogenesis and isomiR levels in tissues or biofluids are often low and heterogeneous^23^.

Mutations in the genes encoding for a handful of RNA binding proteins were found to be linked to increased risk of developing ALS, including Fused in Sarcoma (FUS)^24,25^, and TAR DNA-binding protein 43 (TDP-43)^26^. Cytoplasmic mislocalization of TDP-43 into inclusions and loss of its nuclear function are reported in approximately 97% of patients^27^. Accordingly, cryptic exon splicing impacts dozens of mRNAs^28–30^, some of which may have value as novel biomarkers relevant to ALS therapy development^31,32^. Interestingly, loss of TDP-43 or FUS also leads to dysregulation in isomiR profiles or miRNA biogenesis^33,34^. Furthermore, miRNA biogenesis proteins, Drosha and Dicer, were suggested to be involved in the disease^11,12,35^. Taken together, changes in isomiRs may be directly or indirectly related to ALS. The potential of cell-free isomiRs as biomarkers has been suggested in cancer^36–40^ and in a recent work of Loher et al.^41^. In this work the authors provided profiles and performed analysis of transfer RNA-derived fragments (tRFs), ribosomal RNA-derived fragments (rRFs), Y RNA-derived fragments, and isomiR alterations in ALS plasma of a single cohort. Loher et al. found dozens of small non coding RNAs with diagnostic and prognostic potential. Here, we test the value of isomiRs for ALS prognostication. We profiled cell-free isomiRs in a British cohort of 154 patients with ALS. Using Cox proportional hazard regression^42–44^ and orthogonally machine learning (ML) survival prediction^42,44–47^, we identified 18 isomiR predictors, which were successfully replicated in an independent international cohort of 200 patients. We further demonstrate that isomiR-let-7g-5p.t particularly carries prognostic value, with potential to complement established clinical features and previously described biomarkers, such as miR-181 and NfL. These findings establish isomiR as a novel family of biomarkers for neurodegeneration and highlight their potential as reliable predictors of ALS survival.

## Results

### Study population and baseline characteristics

We conducted our study in two independent cohorts of patients with ALS. Clinical data and blood samples were obtained for 154 patients from the ALS biomarker cohort (UK)^48^, and for 200 patients from the Phenotype Genotype Biomarker (PGB) study^49^. Clinical and demographic cohort characteristics are summarized in **Table 1**. We found the discovery and replication cohorts to be clinically comparable in DeltaFRS (Mann-Whitney P-value = 0.18, **Figure 1A**), baseline ALSFRS-R score (Mann-Whitney P-value = 0.24, **Figure 1B**), site of symptom onset (X^2^ goodness of fit P-value = 0.08, **Figure 1C**), Riluzole treatment (^2^ goodness of fit P-value = 0.34, **Figure 1D**), and C9orf72 repeat expansion status (^2^ goodness of fit P-value = 0.6). Imbalances in patients’ sex distribution (^2^ goodness of fit P-value = 0.009, **Figure 1E**) and age were noticed (t-test P-value ≤ 0.001, **Figure 1F**). The number of censored events were also statistically different between the cohorts (9% vs 54% in UK and PGB, respectively, X^2^ goodness of fit P-value ≤ 0.001, **Figure 1G**).

**Table 1.**
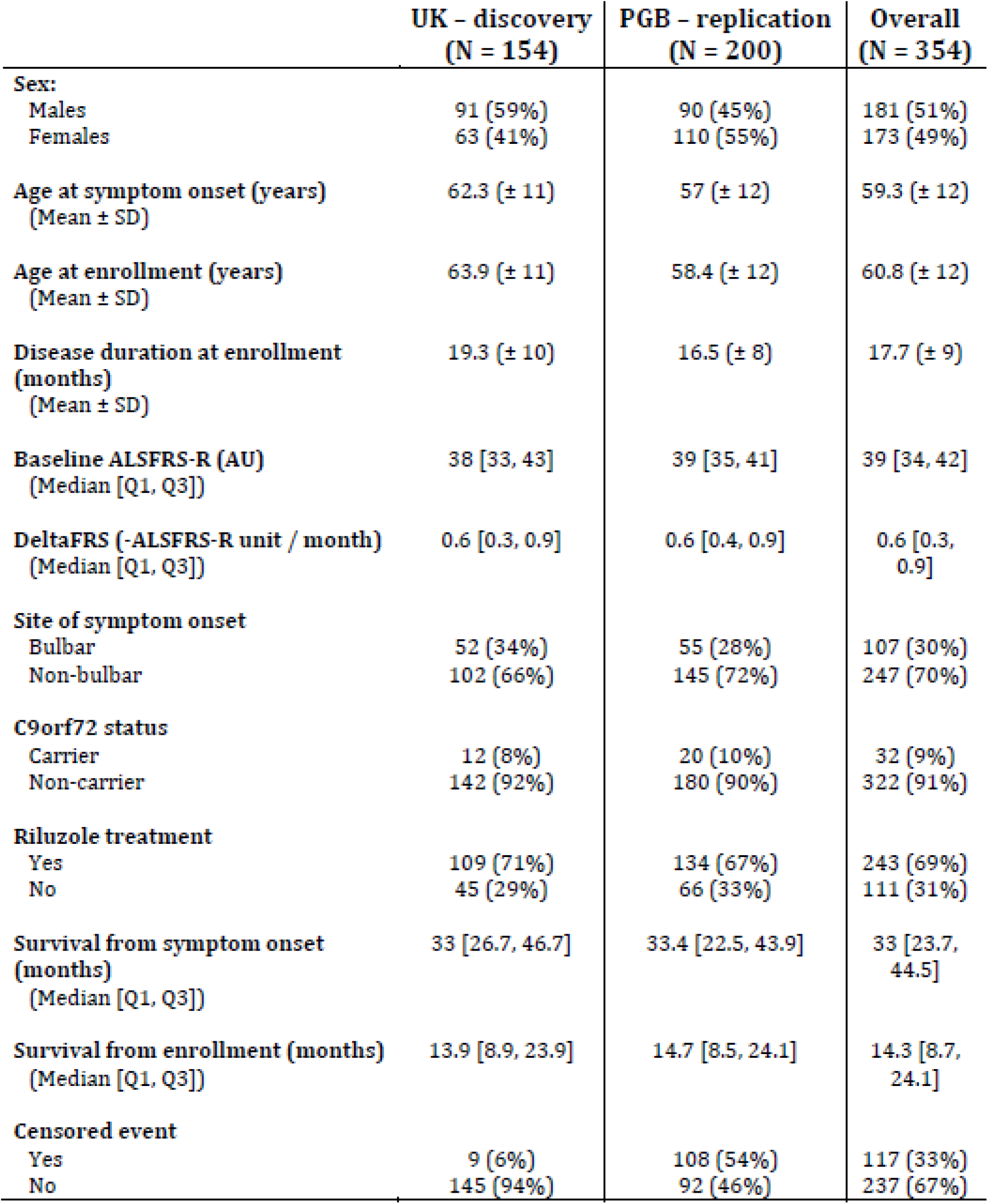
Clinical and demographic description of the study cohorts. Abbreviations: Baseline ALSFRS-R - ALSFRS-R score at baseline assessment. DeltaFRS – change of ALSFRS-R score from symptom onset to baseline assessment divided by months passed. Age at enrollment – age at baseline assessment. Survival from symptom onset / enrollment – time from symptom onset or baseline assessment to death/event censored. Censored event – number and percentage of right-censored cases. SD standard deviation. Q1 - 25th percentile, Q3 - 75th percentile.

|  | UK – discovery<br>(N = 154) | PGB – replication<br>(N = 200) | Overall<br>(N = 354) |
| --- | --- | --- | --- |
| **Sex:** |  |  |  |
| Males | 91 (59%) | 90 (45%) | 181 (51%) |
| Females | 63 (41%) | 110 (55%) | 173 (49%) |
| **Age at symptom onset (years)**<br>(Mean ± SD) | 62.3 (± 11) | 57 (± 12) | 59.3 (± 12) |
| **Age at enrollment (years)**<br>(Mean ± SD) | 63.9 (± 11) | 58.4 (± 12) | 60.8 (± 12) |
| **Disease duration at enrollment (months)**<br>(Mean ± SD) | 19.3 (± 10) | 16.5 (± 8) | 17.7 (± 9) |
| **Baseline ALSFRS-R (AU)**<br>(Median [Q1, Q3]) | 38 [33, 43] | 39 [35, 41] | 39 [34, 42] |
| **DeltaFRS (-ALSFRS-R unit / month)**<br>(Median [Q1, Q3]) | 0.6 [0.3, 0.9] | 0.6 [0.4, 0.9] | 0.6 [0.3, 0.9] |
| **Site of symptom onset** |  |  |  |
| Bulbar | 52 (34%) | 55 (28%) | 107 (30%) |
| Non-bulbar | 102 (66%) | 145 (72%) | 247 (70%) |
| **C9orf72 status** |  |  |  |
| Carrier | 12 (8%) | 20 (10%) | 32 (9%) |
| Non-carrier | 142 (92%) | 180 (90%) | 322 (91%) |
| **Riluzole treatment** |  |  |  |
| Yes | 109 (71%) | 134 (67%) | 243 (69%) |
| No | 45 (29%) | 66 (33%) | 111 (31%) |
| **Survival from symptom onset (months)**<br>(Median [Q1, Q3]) | 33 [26.7, 46.7] | 33.4 [22.5, 43.9] | 33 [23.7, 44.5] |
| **Survival from enrollment (months)**<br>(Median [Q1, Q3]) | 13.9 [8.9, 23.9] | 14.7 [8.5, 24.1] | 14.3 [8.7, 24.1] |
| **Censored event** |  |  |  |
| Yes | 9 (6%) | 108 (54%) | 117 (33%) |
| No | 145 (94%) | 92 (46%) | 237 (67%) |

**Figure 1.**
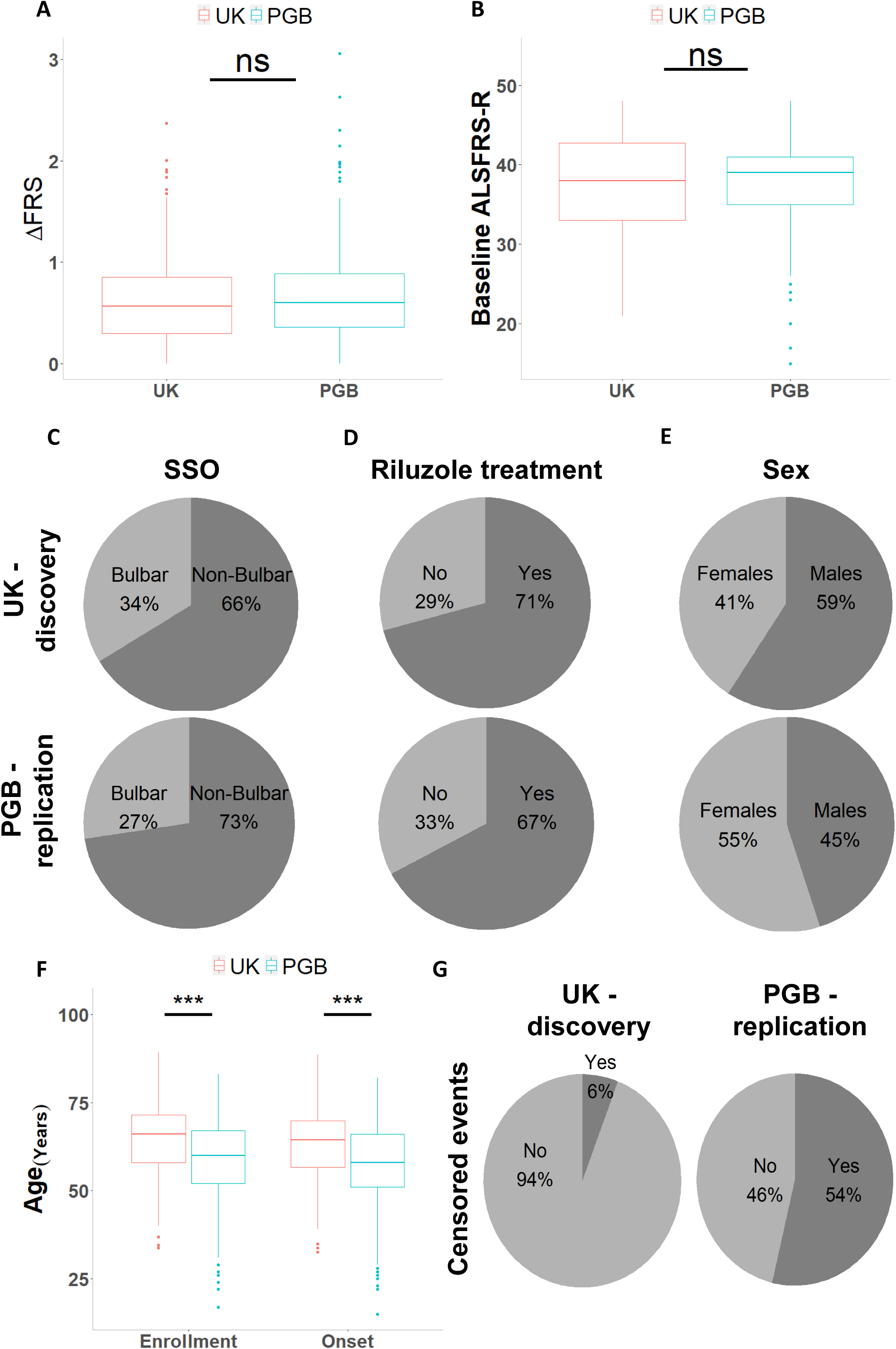
Comparison of the UK discovery and PGB replication cohort metadata. **(A and B)** Boxplot of **(A)** DeltaFRS, measured as the reduction in the ALSFRS-R score per month or **(B)** baseline ALSFRS-R score at study enrollment. **(C – E)** Pie charts depicting **(C)** site of symptom onset (SSO) **(D)** Riluzole treatment, or **(E)** sex **(F)** Boxplots of age at study enrollment and age at symptom onset. **(G)** Pie charts showing the number of censored and deceased events for the UK discovery and the PGB replication cohorts. *** P-value ≤ 0.001, two-sided Student’s t-test.

Our study explores isomiRs, which are sequence isoforms of miRNAs that arise from differences in processing, editing, or modifications of miRNA precursors (**Table S1** for nomenclature; **Figure 2A**), as ALS prognostic biomarkers. Here we use the term “reference miRNA” to refer to the most abundantly expressed (predominant) sequence variant among all isomiRs derived from a given miRNA gene. Noteworthy, the literature refers to a “miRNA” as the collective set of all dozens of annotated isomiRs^20^.

**Figure 2.**
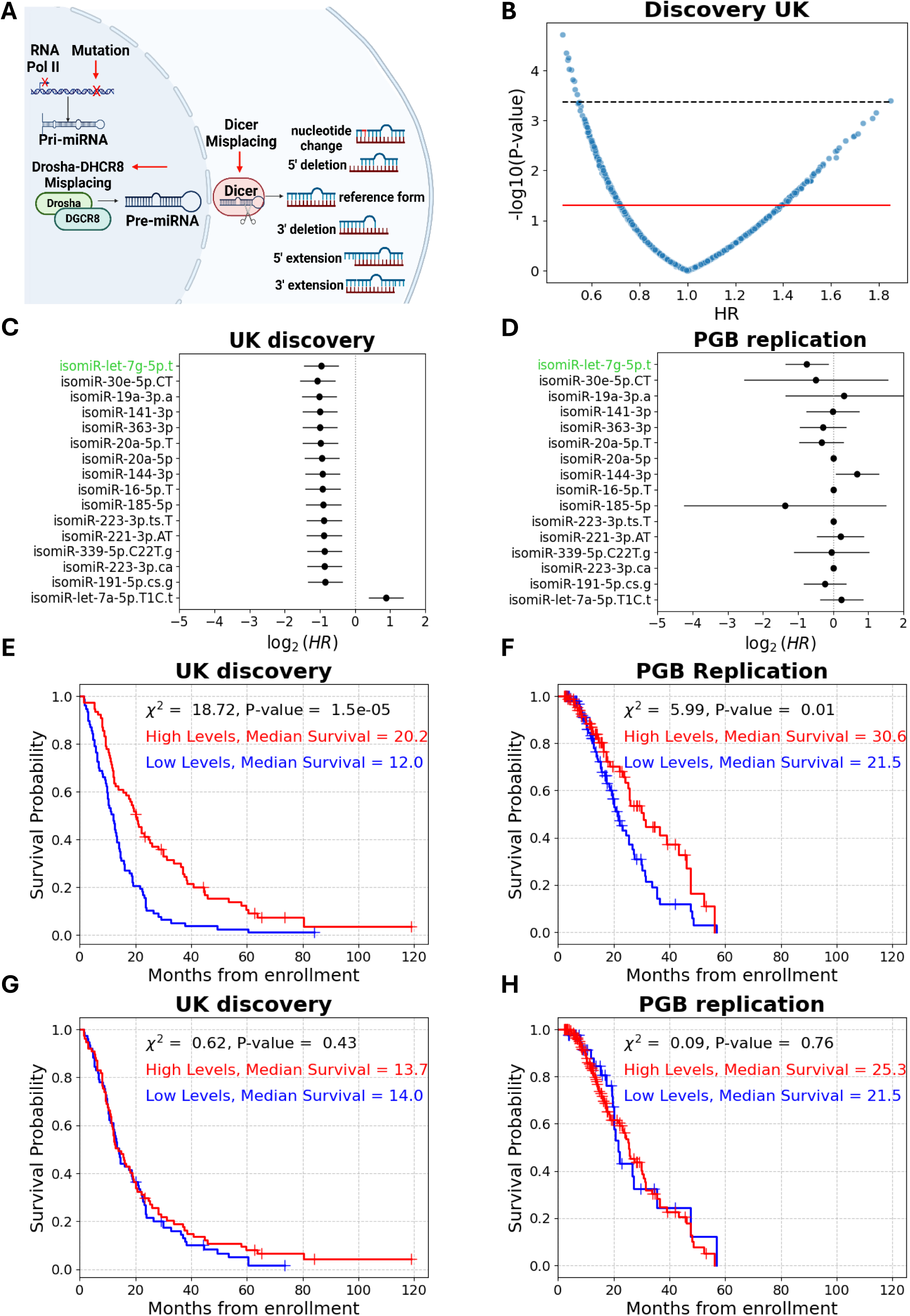
High plasma isomiR-let-7g-5p.t levels predict longer survival in patients with ALS. **(A)** miRNA biogenesis yields several forms of miRNA isoforms (isomiRs). Following transcription, the primary miRNA is cleaved by the nuclear microprocessor, exported as a precursor to the cytoplasm, and further processed by Dicer into a ∼22-nt guide RNA that is loaded onto RISC for target mRNA binding and silencing. Variability in biogenesis, such as misplacement of processing factors, leads to the generation of isomiRs. **(B)** Volcano plot of 1480 plasma isomiRs. Univariate Cox Hazard ratios (HR, x axis) and significance (-log_10_p value; y axis). Red line denotes significance threshold (P-value = 0.05). Only 16 isomiRs remained significant post multiple comparison correction (black line denotes FDR = 5.2e-4). **(C and D)** Forest plots of univariate Cox proportional hazard ratios of the 16 significant isomiRs in UK discovery **(C)** or PGB replication **(D)** cohort (HR ≤ 0.6, or HR = 1.8, P-value ≤ 0.015). Only isomiR-let-7g-5p.t remained significant in both cohorts (denoted by green text). CI denoted by a horizontal line. **(E and F)** Cumulative survival (Kaplan–Meier) curves for isomiR-let-7g-5p.t in the UK discovery **(E)** or PGB **(F)** replication cohort. (UK: 77/77 patients in sub-vs. supra-threshold cohorts; PGB: 104/96 patients in sub-vs. supra-threshold cohorts). **(G and H)** Kaplan–Meier curves for the mature miRNA counterpart, let-7g-5p, in the UK discovery **(G)** or PGB **(H)** replication cohort. (UK: 77/77 patients in sub-vs. supra-threshold sub cohorts; PGB: 40/160 patients in sub-vs. supra-threshold; values used for dichotomization were the same in both cohorts).

Sequencing and data preprocessing were conducted in parallel across 154 and 200 patient samples from the UK and PGB cohorts, respectively. Following noise filtration, 1,480 isomiRs from 287 miRNA genes were co-identified in both the discovery and replication cohorts. The discovery isomiRs study was performed on the UK cohort, whereas the PGB was held out as an unseen, independent replication cohort.

### isomiR-let-7g-5p.t levels differentiate between short and long survival

To identify potential isomiRs associated with patient survival, we dichotomized the 1480 isomiRs that were shared between cohorts, based on their median abundance in the UK discovery cohort. A univariate Cox regression, used to assess the association between isomiR expression and time to death, revealed 290 statistically significant isomiRs (P-value ≤ 0.05), of which 16 remained significant after correction for multiple comparisons (FDR ≤ 0.05, **Figure 2B**). Levels above median of 15 isomiRs were associated with decreased rate of mortality in patients (univariate Cox, Hazard ratios (HR) < 1, 95% confidence interval (CI) = 0.34–0.77, FDR ≤ 0.05; **Figure 2C**), and levels above median of one isomiR were associated with increased rate of mortality (univariate Cox, HR = 1.8, 95% (CI) = 1.32–2.6, FDR ≤ 0.05; **Figure 2C**). Next, we tested these associations in the PGB replication cohort by dichotomizing isomiR levels, using the threshold obtained in the UK discovery cohort. Only levels above median (≥246 unique molecular identifier (UMI) counts) of isomiR-let-7g-5p.t were found to be associated with decreased rate of mortality in the PGB replication cohort (univariate Cox, HR = 0.6, 95% CI = 0.39–0.91, P-value = 0.015; **Figure 2D**). In addition, levels above median of isomiR-let-7g-5p.t were significantly associated with longer survival in the UK discovery cohort (log rank, X² = 18.7, P-value ≤ 0.001, median survival difference = 8.2 months; **Figure 2E**), and in the PGB replication cohort (log rank, X² = 5.99, P-value ≤ 0.01, median survival difference = 9.1 months; **Figure 2F**). Mean isomiR-let-7g-5p.t levels were significantly different in the UK discovery sub cohorts, defined by plasma isomiR-let-7g-5p.t levels that are either higher or lower than the median isomiR-let-7g-5p.t plasma value (351 vs. 176 UMIs in the above- and below-median, Mann-Whitney P-value ≤ 0.001,**Figure S1A**). We found a similar difference in the PGB replication sub cohorts (324 vs. 198 UMIs, Mann-Whitney P-value ≤ 0.001, **Figure S1A**).

Post division by median levels, the sub-cohorts were clinically comparable within each cohort. Age at symptom onset (Mann-Whitney P-value ≥ 0.1, **Figure S1B**), Age at enrollment (Mann-Whitney P-value ≥ 0.08, **Figure S1C**), baseline ALSFRS-R score (Mann-Whitney P-value ≥ 0.27, **Figure S1D**), DeltaFRS (Mann-Whitney P-value ≥ 0.13, **Figure S1E**), sex distribution (X2 goodness of fit P-value ≥ 0.1, **Figure S1F**), site of symptom onset (X2 goodness of fit P-value ≥ 0.2, **Figure S1G**), Riluzole treatment (X2 goodness of fit P-value ≥ 0.15, **Figure S1H**), and C9orf72 repeat expansion status (7 vs 5 and 8 vs 12 carriers in above/below median groups, respectively, in each cohort, X2 goodness of fit P-value ≥ 0.6). Therefore, isomiR-let-7g-5p.t levels stratify patients independently of key clinical variables.

We sought to compare the mortality rate associated with isomiRs and the respective miRNAs, which reflect the aggregate of dozens of expressed isomiRs assigned to a miRNA gene. While isomiR-let-7g-5p.t levels were associated with decreased mortality rate, no cognate miRNA was significantly associated with mortality rate in both cohorts, including let-7g-5p (univariate Cox, HR = 0.87, 1.1; 95% CI = 0.63–1.2, 0.65-1.8; P-value > 0.05 in both, respectively, **Table S2**). Accordingly, plasma levels of cognate miRNA let-7g-5p did not differentiate between short and long survival in the UK discovery or the PGB replication cohorts (log rank X² = 0.63, 0.09; P-value > 0.05 in both, median survival difference = 0.3, 3.8 months, respectively; **Figure 2G, H**).

### IsomiR-let-7g-5p.t is the best prognostication isomiR in two independent ALS cohorts

Although isomiR-let-7g-5p.t emerged as a promising candidate for ALS prognostication, the high dimensionality of our dataset motivated us to explore the predictive performance of isomiR combinations and their potential non-linear interactions using ML, as employed in our previous studies^50^. We sampled at random subsets of 20 isomiRs, with repetition, using bootstrapping, so each isomiR within the pool of 1480 would be included in at least 50 distinct subsets. To mitigate the risk of false positive discoveries inherent in large-scale analyses, the 154 patients of the UK discovery cohort were divided randomly into 10 bins, and a Sequential forward selection (SFS) with 10-fold cross-validation (CV) technique was performed for each of the isomiR subset^51,52^. 84 isomiRs were selected within 90% of the subsets in which they were included after SFS (≥45 out of 50, with Cox regression^42^, random survival forest (RSF)^44^, gradient survival boosting (XGB)^43^**, Figure S2A**). Of the three orthogonal survival models, XGB demonstrated the highest mean concordance index score (C-index) across the 84 isomiRs (**Figure S2B**).

Next, we modeled survival of the 84 isomiRs by XGB. IsomiRs were included if their subsets displayed a C-index higher than a threshold that was defined empirically by the elbow method^53^ (> C-index of 0.74). Reassuringly, isomiR-let-7g-5p.t was detected, along with 17 other isomiRs: isomiR-let-7e-5p.G9C, isomiR-let-7f-5p.T16C.t, isomiR-let-7i-5p.G5A, isomiR-130b-5p.T, isomiR-142-3p.a, isomiR-451a.T.A, isomiR-144-3p, isomiR-146a-5p.T22A, isomiR-151a-3p.A9G, isomiR-25-3p.A22T, isomiR-30d-5p.A20G.CT, isomiR-425-5p.as, isomiR-432-5p, isomiR-486-5p.C, isomiR-486-5p.T9G.T, isomiR-92a-3p.T1C, isomiR-92a-3p.T22C (**Figure S2C**).

After hyperparameter tuning with 5-fold CV (Supplementary Methods), a model based on these 18 isomiRs demonstrated strong prognostic capacity across patient survival duration of 84 months (time-dependent area under the curve (AUC(t)) values of 0.7 to 0.95; average 0.78; C-index = 0.7; survival duration 2 - 85 months; **Figure 3A**), but survival prediction was less effective in the PGB replication cohort (AUC(t) 0.49-0.9, average 0.62; C-index = 0.55; survival duration 3–54 months; **Figure 3A**). Notably, prediction was better for patients with longer survival times. Additionally, miRNA and reference miRNA displayed poor performance over time (**Figure 3B, C**).

**Figure 3.**
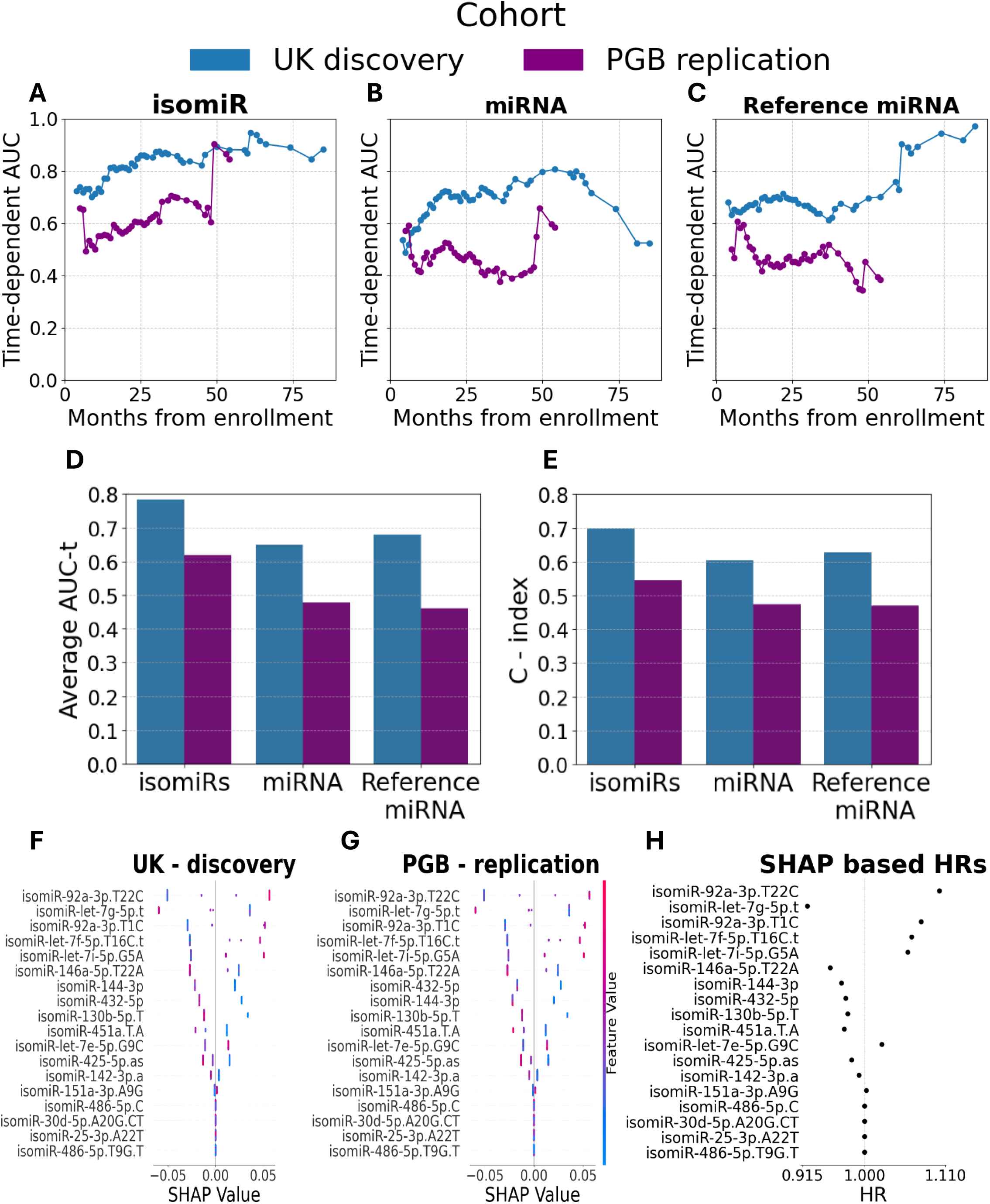
IsomiR based survival prediction in two independent cohorts. **(A-C)** Plots of AUC (y axis) in the UK discovery (blue) or PGB replication (purple) cohorts for patient survival, as a function of duration from enrolment (x axis) for isomiR **(A)**, cognate miRNA **(B)**, or reference miRNA **(C)**. **(D and E)** Bar graphs of **(D)** average AUC across time (47/42 time points for patients from the UK discovery or PGB replication, respectively), or **(E)** C-index for a model based on 18 selected isomiRs. **(F and G)** SHAP analysis of the contribution of 18 isomiRs to risk prediction in the **(F)** UK discovery or **(G)** PGB replication cohorts. Range of isomiR levels (low–blue to high–red). Positive or negative SHAP values reveal higher or lower effect size of isomiR on the mortality risk (x axis), respectively. **(H)** Plot of HR, calculated on the basis of XGB SHAP values according to Sundrani & Lu^54^.

We demonstrate higher performance with the model trained on the 18 selected isomiRs relative to models trained on the collective set of all annotated isomiRs (the ”miRNA”, average AUC(t) = 0.64/0.53, C-index= 0.59/0.52 in the UK discovery and PGB replication cohorts, respectively) or the most abundant isomiR (“reference miRNAs” average AUC(t)= 0.68/0.46, and C-index = 0.63/0.47, **Figure 3D, E**).

To identify the isomiRs most valuable for survival prediction, we calculated Shapley Additive Explanations (SHAP) for the model average risk function. High levels of isomiR-let-7g-5p.t were associated with reduced risk of death and longer survival (**Figure 3F**). In contrast, high levels of isomiR-92a-3p.T22C, isomiR-92a-3p.T1C, isomiR-let-7f-5p.T16C.t, and isomiR-let-7i-5p.G5A associate with higher risk and shorter survival. A similar SHAP analysis of the PGB replication cohort was highly correlated, highlighting the model’s robustness (**Figure 3G**). We then derived the HRs from SHAP values, as previously described^54^. IsomiR-92a-3p.T22C and isomiR-let-7g-5p.t levels were associated the most with increased / decreased mortality, respectively (**Figure 3H**).

As isomiR expression can vary across cohorts, we assessed abundance and inter-cohort correlation to evaluate biological reproducibility. Among the 18 selected isomiRs, mean expression ranged from 4–9 UMIs (UK cohort), and 5–14 UMIs (PGB cohort) and the detection rates were comparable between cohorts (**Figure S3A, B**). IsomiR-142-3p.a had the highest expression (UK = 974_±_833 UMIs, PGB = 1984_±_945 UMIs), followed by isomiR-let-7g-5p.t (UK discovery cohort: 264 UMIs ± 129; PGB replication cohort: 259 UMIs ± 94). IsomiR abundance was well correlated between cohorts (r = 0.74, P = 8.1e−28, **Figure S3C, D**) but no correlation was noted between the abundance of isomiRs and their “cognate miRNA”, nor with the “reference miRNA” (**Table S3**).

### Lower isomiR-let-7g-5p.t levels in ALS blood relative to non-neurodegeneration controls

Having found that isomiR-let-7g-5p.t holds prognostic value for ALS in two orthogonal approaches, we compared its levels between patients and controls. To do so, we reanalyzed plasma samples from two independent control cohorts from Queen Mary hospital (QMUL - CTL) and University College London (UCL - CTL). IsomiR-let-7g-5p.t levels were comparable across the two ALS cohorts within the same sub cohort (mean difference ≤ 27 UMI, Tukey honestly significant difference [HSD] P-value ≥ 0.99, **Figure S4**), and although variable across control cohorts (mean difference = 324, Tukey HSD P-value ≤ 0.001), we observed significantly lower levels in the below-median ALS sub cohorts compared to both controls and the above-median ALS sub cohorts (mean difference ≥ 369 UMI and mean difference ≥ 522 UMI, respectively, Tukey HSD P-value ≤ 0.001, **Figure S4**). Therefore, isomiR-let-7g-5p.t is also of potential diagnostic value.

Lastly, after examining isomiR-let-7g-5p.t biomarker potential, we tested if there was a change in the isomiR’s predicted targets compared to its reference miRNA. We retrieved the predicted targets of miRNA let-7g-5p from miRCarta^55^, subjected them to a miRNA-mRNA hybridization prediction algorithm^56^ and compared to the reference miRNA let-7g-5p and isomiR-let-7g-5p.t. Out of potential ∼4000 mRNA targets, 3189 had at least one significant hybridization site with either the reference miRNA let-7g-5p or isomiR-let-7g-5p.t. 358 out of 3189 mRNA targets were predicted to lose at least one hybridization site, and 43 were predicted to gain at least one such isomiR-let-7g-5p.t site (Wilcoxon p-value = 2.4e-47, rank-biserial correlation effect size = 0.67). Enrichment for targets associated with ALS was not noticed.

### Prognostic Value of Plasma isomiR-let-7g-5p.t in ALS Is Comparable to NfL

As we previously reported^17,57^, high (above median) blood NfL levels were significantly associated with shorter survival in the UK discovery cohort (log-rank, X² = 9.4, P-value = 2e-3, median survival difference = 7.1 months; **Figure 4A**), and this was consistent in the PGB replication cohort, when NfL dichotomized according to the median plasma value in the UK cohort (log rank, X² = 17.7, P-value = 2.7e-5, median survival difference = 13.7 months; **Figure 4B**). Above-median NfL levels were associated with an increased mortality rate in both cohorts (UK discovery cohort: univariate Cox, HR = 1.7, 95% CI = 1.2-2.3, P-value = 2.4e-4, C-index = 0.56 ; PGB replication cohort Cox HR = 2.4, 95% CI 1.6-3.7, P-value = 4.5e-6, C-index = 0.63; **Figure 4C, D**). In contrast, higher levels of seven (out of eighteen selected by ML, **Figure S3C**) isomiRs were associated with a decreased mortality rate (univariate Cox, HR < 0.72, 95% CI = 0.37-0.99, P-value ≤ 0.05, C-index > 0.54; **Figure 4C**). However, only isomiR-let-7g-5p.t retained significant HR in univariate Cox regression in the PGB replication cohort (UK discovery cohort: HR = 0.58, 95% CI = 0.41-0.81, P-value = 1.4e-3, C-index = 0.57; PGB replication cohort: univariate Cox, HR = 0.6, 95% CI = 0.4-0.9, P-value = 0.015, C-index = 0.55; **Figure 4C, D**). Accordingly, isomiR-let-7g-5p.t was the only isomiR that significantly distinguishes short and long survival times in both cohorts, using the log-rank test (**Figures S5A, B**).

**Figure 4.**
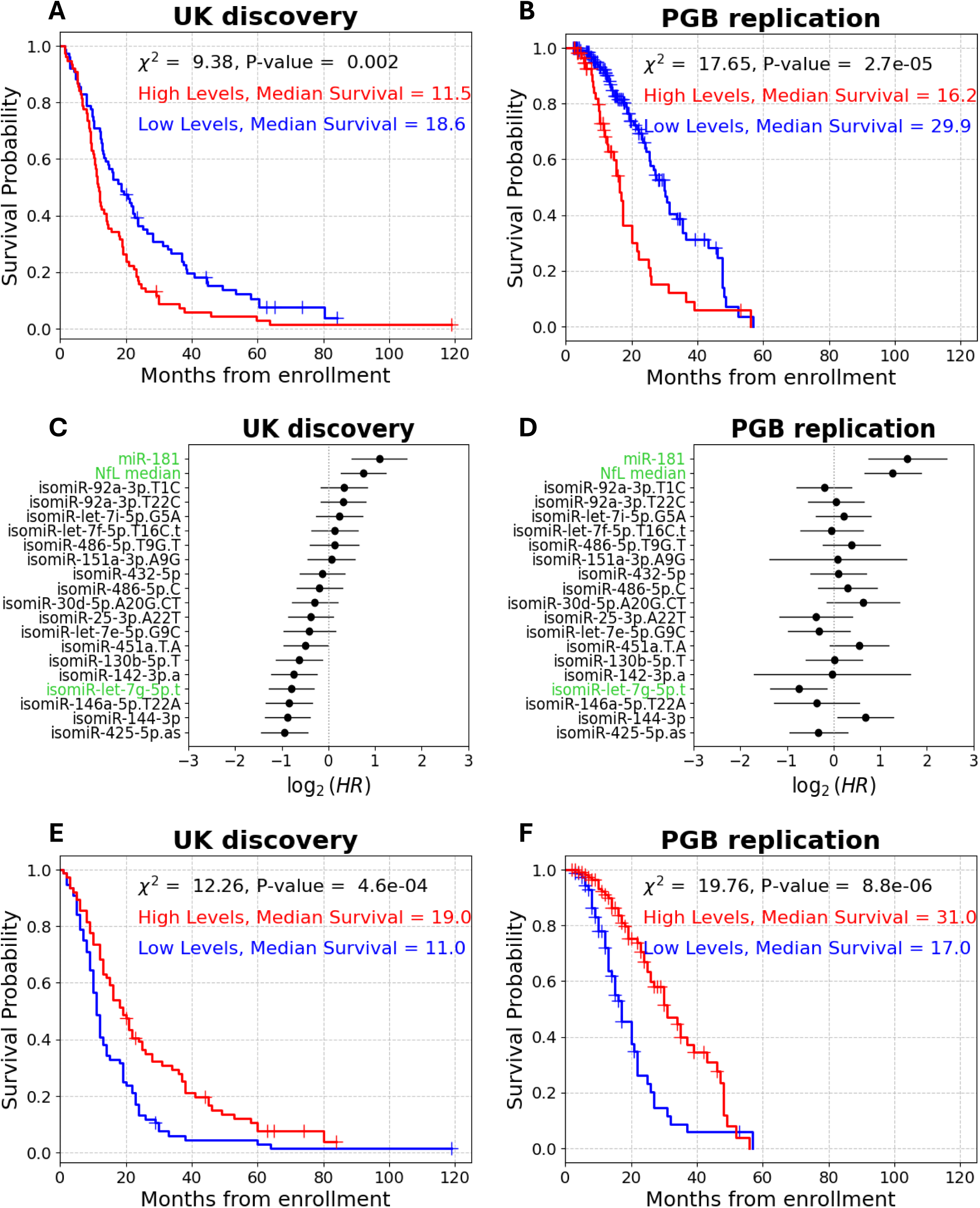
IsomiR-let-7g-5p.t-based prognostication compared to NfL. **(A and B)** Cumulative survival (Kaplan–Meier) curves for NfL plasma levels (76/76 sub-threshold (blue) /supra-threshold (red) patients, respectively, from enrollment in the UK discovery cohort **(A)** and **(B).** NfL serum levels (140/60 sub-threshold (blue) /supra-threshold (red) patients, respectively) in the PGB replication cohort **(B)**. **(C and D)** Forest plots of univariate Cox proportional hazard ratios of 18 isomiRs, NfL and miR-181 in **(C)** the UK discovery cohort or **(D)** the PGB replication cohort. NfL, miR-181 and isomiR-let-7g-5p.t are highlighted in green. CI denoted by horizontal line. **(E and F)** Cumulative survival (Kaplan–Meier) curves for isomiR-let-7g-5p.t/NfL ratio (76/76 sub-threshold (blue) / supra-threshold (red) patients, respectively) from enrollment in **(E)** the UK discovery cohorts of **(F)** the PGB replication cohort (53/147 sub-threshold (blue) / supra-threshold (red) patients, respectively)

In multivariate Cox analysis, NfL remained significantly associated with increased mortality rate in both cohorts (UK discovery HR = 1.7, 95% CI = 1.2-2, P-value = 6e-4; PGB replication HR = 3.3, 95% CI = 2.4-5.3, P-value = 2e-7), whereas isomiR-let-7g-5p.t was significant only in the PGB cohort (multivariate Cox, HR = 0.52, 95% CI = 0.32-0.86, P-value = 0.01; **Figure S6A, B**). Noteworthy, isomiRs modestly improved the prognostic capacity, compared to a model which only includes NfL, in both cohorts (C-index = 0.67/0.72 in UK discovery or PGB replication, respectively, compared to 0.56/0.63 for NfL alone).

When considering only isomiR-let-7g-5p.t and NfL together, isomiR-let-7g-5p.t showed a stronger association with risk of mortality than NfL in the UK discovery cohort (HR of NfL in a multivariate Cox model = 1.64, 95% CI = 1.2-2.3, P-value = 3.9e-4; HR of isomiR-let-7g-5p.t in a multivariate Cox model = 0.49, 95% CI = 0.35-0.69, P-value = 3.8e-6; **Figure S6C**). Together, NfL and isomiR-let-7g-5p.t yield better prognostication than either marker alone (multivariate Cox C-index = 0.63 vs. C-index = 0.56 for NfL alone or C-index = 0.59 for isomiR-let-7g-5p.t alone). In the PGB cohort, NfL had a stronger association with mortality rate (NfL multivariate Cox, HR = 3, 95% CI = 1.9-4.6, P-value = 1e-7; isomiR-let-7g-5p.t multivariate Cox, HR = 0.46, 95% CI = 0.29-0.7, P-value = 4.8e-5; **Figure S6D**). Nonetheless, both markers together improved survival prediction compared to either one individually (multivariate Cox C-index = 0.67, for NfL alone C-index = 0.63, for isomiR-let-7g-5p.t alone C-index = 0.56). Moreover, when combined with relevant clinical features (baseline ALSFRS-R, site of symptom onset, age at onset), only isomiR-let-7g-5p.t (UK discovery multivariate Cox, HR = 0.53, 95% CI = 0.38-0.75, P-value = 3e-5; PGB replication multivariate Cox, HR = 0.52, 95% CI = 0.33-0.83, P-value =6.5e-3) and baseline ALSFRS-R (UK discovery multivariate Cox, HR = 0.97, 95% CI = 0.94-0.99, P-value = 2.6e-3; PGB replication multivariate Cox, HR = 0.96, 95% CI = 0.92-0.99, P-value = 0.013) were significantly associated with mortality risk in both cohorts (**Figure S6E, F**). In contrast, NfL levels association with mortality were only approaching significance in the UK discovery cohort (multivariate Cox, HR = 1.4, 95% CI = 0.97-1.94, P-value = 0.08; **Figure S6E**), while being significantly associated with mortality in the PGB cohort, with an effect size larger than that of isomiR-let-7g-5p.t (multivariate Cox, HR = 2.5, 95% CI = 1.5-4, P-value ≤ 0.001; **Figure S6F**).

Since above-median levels of isomiR-let-7g-5p.t and NfL were associated with opposing effects on mortality risk, we next assessed joint prognostic capacity, by calculating the ratio between isomiR-let-7g-5p.t and NfL levels. High values of this ratio slightly improved distinction between short and long survival compared to NfL alone (UK discovery log-rank, X² = 11.8, P-value = 5.9e-4, median survival difference = 8 months; PGB replication log-rank, X² = 19.8, P-value = 8.8e-6, median survival difference = 14 months; **Figures 4E, F**). Notably, the ratio showed a stronger inverse association with mortality risk compared to either marker alone, and improved overall prognosis (UK discovery univariate Cox, HR = 0.56, 95% CI = 0.4-0.78, P-value = 6.2e-4, C-index = 0.58; PGB replication univariate Cox, HR = 0.4, 95% CI = 0.26-0.6, P-value = 1.8e-5, C-index =0.64 ;**Figure S6G**). Taken together, these results suggest that isomiR-let-7g-5p.t performs comparably to NfL, and that combining isomiR-let-7g-5p.t with NfL improved predictive performance, at least as reflected by the C-index. These findings highlight the value of isomiR-let-7g-5p.t as a potential prognostic biomarker in ALS.

### Prognostic Value of Plasma isomiR-let-7g-5p.t in ALS Is Comparable to miR-181

The UK cohort was a subset of the cohort where we originally discovered miR-181, a multiplication of sister miRNAs miR-181a-5p and miR-181b-5p, as a prognostic marker^17^. A univariate Cox analysis revealed that high miR-181 levels were associated with a >2.1-fold increased risk of mortality, again demonstrating that isomiR-let-7g-5p.t has an equivalent prognostic capacity to previously published markers (UK discovery cohort HR = 2.1, 95% CI = 1.4–3.2, P-value ≤ 0.001, C-index = 0.58; PGB replication cohort HR = 3.0, 95% CI = 1.7–5.4, P-value ≤ 0.001, C-index = 0.55; **Figure 4C, D**).

miR-181 levels distinguished between short and long survivors in the UK cohort (log rank on dichotomized miR-181 levels: X² = 13.7, P-value = 2.1e-4, median survival difference = 7.2 months; **Figure 5A**), and also in the PGB cohort (log rank: X² = 15.1, P-value = 1e-4, median survival difference = 9.7 months; **Figure 5B**). Notably, isomiR-let-7g-5p.t was the only isomiR to remain statistically significant in both cohorts, in a combined analysis of miR-181 and the 18 isomiR chosen by ML (UK discovery multivariate Cox, HR = 0.63, 95% CI = 0.4-0.98, P-value = 0.04; PGB replication multivariate Cox, HR = 0.57, 95% CI = 0.36-0.9, P-value = 0.017; **Figure 5C**, D). To assess joint prognostic capacity of isomiR-let-7g-5p along with miR-181, we tested isomiR-let-7g-5p / miR-181 ratio capacity to distinguish between short and long survival. We found a significant association of the ratio levels to patient survival in both cohorts (UK discovery log rank: X² = 20.6, P-value = 5.6e-6, median survival difference = 8 months; PGB replication log rank: X² = 7, P-value = 8e-3, median survival difference = 6 months; **Figures 5E**, F). Moreover, the HR of isomiR-let-7g-5p / miR-181 ratio was 0.38 in the discovery and 0.44 in the replication cohort (UK discovery univariate Cox, 95% CI = 0.19–0.77, C-index = 0.58, P-value = 8e-6; PGB replication univariate Cox, 95% CI = 0.18-0.8, C-index = 0.53, P-value = 0.01, Figure S6G). Taken together, these results highlight the potential of isomiR-let-7g-5p.t as a robust biomarker for ALS survival.

**Figure 5.**
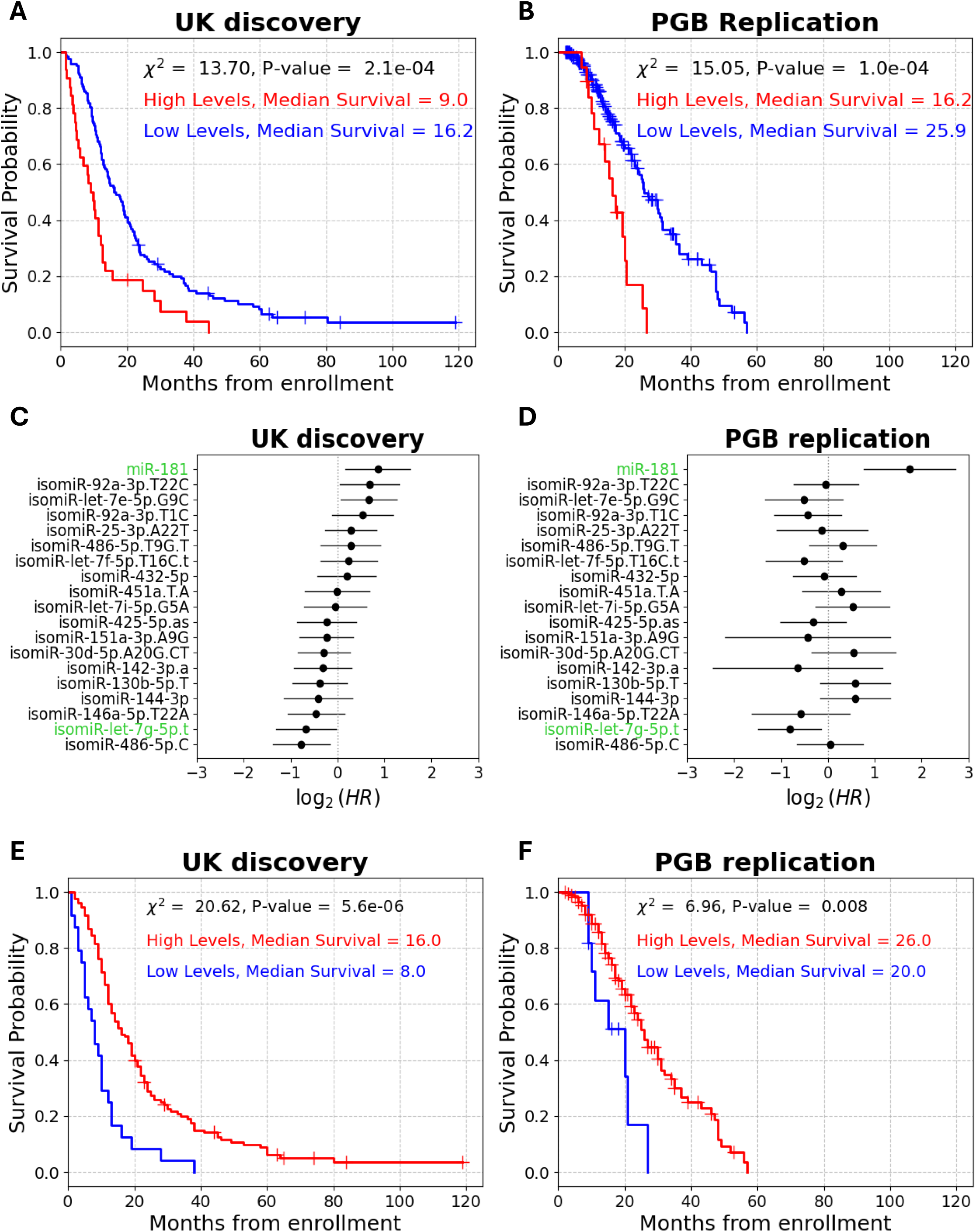
isomiR-let-7g-5p.t-based prognostication compared to miR-181. **(A and B)** Cumulative survival (Kaplan–Meier) for miR-181 (122/32 sub-threshold (blue) / supra-threshold (red) patients, respectively) in the UK discovery **(A)** or PGB replication (181 /19 patients sub-threshold (blue) / supra-threshold (red) patients, respectively, **B)** cohorts. **(C and D)** Forest plots of multivariate Cox proportional hazard ratios of 18 isomiRs and miR-181 in the UK discovery **(C)** or the PGB replication **(D)** cohorts. IsomiR-let-7g-5p.t and miR-181 are highlighted in green. CI denoted by horizontal line. **(E and F)** Cumulative survival (Kaplan–Meier) for isomiR-let-7g-5p.t/miR-181 ratio (24/130 sub-threshold (blue) / supra-threshold (red) patients, respectively) from enrollment in the **(E)** UK discovery cohort or **(F)** PGB replication cohort (11/189 sub-threshold (blue) / supra-threshold (red) patients, respectively)

## Discussion

IsomiRs display small variations of either sequence insertion, deletion, or single nucleotide polymorphism that distinguish them from the reference miRNA form. While miRNAs are extensively studied in ALS^12,14–17,58–67^, we are first to demonstrate the value of isomiRs as biomarkers for neurodegeneration in two large, independent cohorts. Among 1480 studied isomiRs, isomiR-let-7g-5p.t emerges as the most promising prognostic biomarker. High plasma levels of isomiR-let-7g-5p.t are significantly associated with lower mortality rate and longer survival. This effect was not explained by clinical characteristics, suggesting that the information encompassed in isomiR-let-7g-5p.t is not directly associated with a single clinical feature. IsomiR-let-7g-5p.t levels were reduced in two ALS cohorts relative to two control cohorts, suggesting diagnostic potential. Higher plasma levels of isomiR-let-7g-5p.t are comparable but reciprocal to miR-181 and NfL, which we previously associated with ALS prognosis^9,17,57^. Additionally, the incorporation of isomiR-let-7g-5p.t to either miR-181 or NfL enhances their prognostic value.

Dysregulation of miRNAs in ALS has previously been demonstrated in mice and humans^11,15,16^. Numerous studies have proposed miRNA utility as potential biomarkers for prognosis for ALS ^7,61,65^. However, isomiRs have been overlooked in part due to their sequence heterogeneity and their low abundance^23^. Nevertheless, our work, the work of Loher et al.^41^, and notable dysregulation of RNA-binding proteins such as TDP-43, DICER, DROSHA, or FUS in ALS^11,12,34,68,69^ should prompt additional studies of isomiRs in ALS. In addition, it is possible that isomiRs play a role in other neurodegenerative disorders, with implications ranging from basic research, to clinical applications.

Our analysis found that isomiR-let-7g-5p.t accounts for approximately 40% of the reads of let-7g-5p in the plasma and perhaps predominates the let-7g-5p repertoire. In this context, microRNA let-7g-5p has previously been reported to be lowered in abundance in the blood of patients with sporadic ALS (sALS)^65^, and in patients with frontotemporal dementia (FTD) due to a *C9orf72* repeat expansion, compared to healthy controls^70^. Loher et al^41^ reported hundreds of differentially abundant isomiRs in ALS patient plasma compared to controls, with roughly equal numbers up- and down-regulated. 53 out of 56 Let-7 family isomiRs were elevated in ALS. The association of reduced plasma isomiR-let-7g-5p.t levels to severe disease, observed in our study, suggests a distinct behavior not captured by Loher et al^41^. Let-7g-5p blood levels are also decreased in individuals with Alzheimer’s disease (AD) or mild cognitive impairment (MCI)^71–78^, and correlate with AD severity^71^. In addition, let-7g-5p was found to be downregulated in brain samples of Huntington patients compared to controls^79^. This could reflect a specific decrease that is mediated, at least in part, via isomiR-let-7g-5p.t.

Members of the let-7 miRNA family have been repeatedly linked to neurodegenerative diseases. In ALS, let-7d and let-7f are reduced in patient plasma, while let-7b is elevated in leukocytes. In AD, several let-7 members, including let-7b, let-7d, let-7e, and let-7g, are upregulated in patient biofluids or neuronal vesicles, with let-7b also correlating with disease progression. In Parkinson’s disease, let-7a and let-7f are decreased in plasma, whereas let-7g-3p is increased in CSF-derived vesicles^66,80–90^. Together, these findings underscore the relevance of the let-7 family in neurodegeneration and reinforce the significance of isomiR-let-7g-5p.t as a prognostic marker in ALS.

Seventeen other isomiRs displayed potential relevance in our analysis, though to a weaker extent. Some of the miRNAs associated with these isomiRs (miR-451, miR-144-3p, and miR-486-5p) are linked to oxidative stress and inflammation^91–99^, which potentially play a role in neurodegeneration. However, because we analyze the isomiRs in plasma, the tissue expression origin is unclear.

The mechanisms underlying isomiR levels, including isomiR-let-7g-5p.t, and their functional role in disease progression is yet to be fully elucidated. The loss of a single nucleotide at the 3′ end occurs outside the seed region and has limited impact on target specificity^100–103^. Accordingly, using RNA22 v2^56^, we found that a subset of predicted targets might interact differently with isomiR-let-7g-5p.t relative to let-7g-5p, but no enrichment was observed for ALS-related pathways.

Together, our results highlight the potential of isomiR-let-7g-5p.t as a prognostic biomarker in ALS, potentially offering comparable value to clinical features, miR-181, and NfL. Moreover, isomiR-let-7g-5p.t enhances effective prognostication in conjunction with other markers. These findings warrant further exploration of the utility of isomiR-let-7g-5p.t and other isomiRs in neurodegenerative diseases.

### Limitations of Study

Despite the promising prognostic potential of isomiRs in ALS, several limitations must be noted: some uncertainty remains about the reference miRNA due to high sequence similarity within the broad family of Let-7 miRNAs. Future advances in sequencing technologies and alignment algorithms may improve isomiR resolution and assignment. We did not find any influence for sex on the results of the study. Larger and more diverse population cohorts would strengthen the generalizability of the findings and help assess the robustness of isomiRs across different ALS subtypes, sex, or disease stage. Last, given the availability and strong prognostic performance of NfL, the practical utility of isomiRs as standalone biomarkers may be limited until an effective way to measure them is developed. These limitations highlight the need for continued research on isomiRs in ALS.

## Data Availability

The newly generated transcriptomic small RNA-seq datasets have been uploaded to GEO: https://www.omicsdi.org/dataset/geo/GSE298532 for ALS patients and GEO: https://www.omicsdi.org/dataset/geo/GSE307456 for controls. Additional data including clinical characteristics, NfL quantification, corrected transcriptomic data, feature level binarization, univariate Cox results, and ML feature selection results have been uploaded to Mendeley data: https://doi.org/10.17632/2xxw2f28p6.2). The custom code generated in this project, including data preprocessing, analysis, and plotting, has been uploaded to Mendeley data: https://doi.org/10.17632/2xxw2f28p6.2). Any additional information required to reanalyze the data reported in this work paper is available from the lead Contact upon request.

## Acknowledgments

Eran Hornstein is the Mondry Family Professorial Chair and Head of the Nella and Leon Benoziyo Centre for Neurological Diseases and of the Andi and Larry Wolfe Centre for Neuroimmunology and Neuromodulation. We acknowledge participants and families for their contributions. We thank ALS biomarker cohort study (UK) and the CReATe consortium’s Phenotype-Genotype biomarker (PGB) study co-workers for their contributions to biobanking, genomics, biorepository, project management, and data management. We thank Crown Genomics Team of Nancy and Stephen Grand Israel National Center for Personalized Medicine for small RNA sequencing. The authors thank Dr. Isidore Rigoutsos and Phillipe Loher (Thomas Jefferson University), for assistance with the revised manuscript. This study was funded by several programs. Eran Hornstein lab is funded by the Andi and Larry Wolfe Centre for Neuroimmunology and Neuromodulation; the Binational Science Foundation (BSF); Association Française Contre les Myopathies (AFM grants 24882, 28680); Muscular Dystrophy Association (MDA United States grant 1280000), Target ALS, Israel Science Foundation (ISF 3497/21, 424/22, 3643/26), ALS Canada, ALS Association (United States), Minna-James-Heineman Stiftung through, Minerva Foundation, with funding from the Federal German Ministry for Education and Research, Robert Packard Center for ALS Research at Johns Hopkins, McGill University, EU - ERA-Net, Radala Foundation for ALS Research. Additional support generously provided by the Kekst Family Institute for Medical Genetics. Weizmann SABRA - Yeda-Sela - WRC Program, the Estate of Emile Mimran, and The Maurice and Vivienne Wohl Biology Endowment. Nella and Leon Benoziyo Center for Neurological Diseases, Goldhirsh-Yellin Foundation. Dr. Sydney Brenner and friends, Weizmann - Center for Research on Neurodegeneration. Redhill Foundation – Sam and Jean Rothberg Charitable Trust Dr. Dvora and Haim Teitelbaum Endowment Fund, and Weizmann Institute Knell Family Institute for Artificial Intelligence. The CReATe Consortium (U54NS092091) is part of the NIH Rare Diseases Clinical Research Network (RDCRN), an initiative of the Office of Rare Diseases Research (ORDR), NCATS. This Consortium is funded through a collaboration between NCATS and the NINDS. This work was also supported by a Clinical Trial Readiness grant (U01NS107027) from NINDS and by a grant from the ALS Association to support the CReATe Biorepository (grant ID 16-TACL-242).

## Resource availability

### Lead contact

Further information and requests for resources should be directed to and will be fulfilled by the lead contact, Eran Hornstein

### Materials availability

This study did not generate new unique reagents.

### Data and code availability

- The newly generated transcriptomic small RNA-seq datasets have been uploaded to GEO: https://www.omicsdi.org/dataset/geo/GSE298532 for ALS patients and GEO: https://www.omicsdi.org/dataset/geo/GSE307456 for controls. Additional data including clinical characteristics, NfL quantification, corrected transcriptomic data, feature level binarization, univariate Cox results, and ML feature selection results have been uploaded to Mendeley data: https://doi.org/10.17632/2xxw2f28p6.2).
- The custom code generated in this project, including data preprocessing, analysis, and plotting, has been uploaded to Mendeley data: https://doi.org/10.17632/2xxw2f28p6.2).
- Any additional information required to reanalyze the data reported in this work paper is available from the lead Contact upon request.

## Author contributions

E.H., Y.C. and I.M. oversaw the conceptualization of this study, M.B., A.M. and J.W. recruited participants and collected clinical data and biofluid samples. E.H., Y.C., I.M. & I.S. designed the study and had unrestricted access to all data. Y.C. and I.M. oversaw molecular data collection. Y.C., I.M. and I.S. performed, and replicated the statistical analyses with feedback from M.B. and J.W. E.H. oversaw all facets of the study. Y.C., I.M, Y.M.D, M.B, A.M. and E.H participated in drafting the manuscript, and all authors approved the final draft, and take full responsibility for its content, including the accuracy of the data and the fidelity of the trial to the registered protocol and its statistical analysis.

## STARMETHODS

### Experimental model and subject details

#### Human samples

##### Study Cohorts

Two ALS patient cohorts were included.

###### ALS biomarker study (London, UK)

This cohort consists of 154 British patients (REC Reference: 09/H0703/27, IRAS ID: 47352). Patients with neuroinflammatory or neurodegenerative disorders, recent injuries, systemic or organ-specific autoimmune disorders, or recent treatment with steroids, immunosuppressants, or immunoglobulins were excluded. Ethics approval was obtained from East London and the City Research Ethics Committee 1.

###### PGB study subset (CReATe Consortium)

This cohort included 200 patients recruited from 12 U.S. sites and one South African site, as part of the Clinical Research in ALS and Related Disorders for Therapeutic Development (CReATe) Consortium (registered at clinicaltrials.gov NCT02327845) ^9,49^ Cognitive impairment (ALSci) or behavioral impairment (ALSbi) were determined by applying the revised Strong criteria^104^ to Edinburgh Cognitive and Behavioral ALS Screen (ECAS^105^) data. Ethics approval was granted by the University of Miami central IRB (Internal Protocol ID #20160603), and by the University of Cape Town Health Sciences Human Research Ethics Committee in South Africa (REF number 165/2017).

Two control cohorts were analyzed.

###### ALS biomarker study controls

Ninety-nine adult individuals (72 females and 27 males, mean age 50.6 ± 13.2 years) recruited at Queen Mary Hospital, London (ethics approval 09/H0703/27).

###### UCL control cohort

Fifty-six adult individuals (30 females and 26 males, mean age 61 ± 13.3 years) from a longitudinal FTD cohort study at University College London (ethics approval 15/0805).

Controls were typically spouses or relatives of patients that were screened to confirm absence of neurological or neurodegenerative diseases.

All Patients were diagnosed with ALS by experienced ALS neurologists. Inclusion criteria to address a “trial-like” conditions were as follows: time from symptom onset to first blood draw shorter than 36 months, a baseline slow vital capacity (SVC) equal to or above 50%, and at least two study visits with measurement of the revised amyotrophic lateral sclerosis functional rating scale (ALSFRS-R)^106^. All participants provided written consent (or verbal permission for a carer to sign on their behalf) to be enrolled in the ALS biomarker study if they met inclusion criteria. Participants information on sex, and age was self-reported. Information on gender, race, ethnicity and socioeconomic status was not collected. All patients were de-identified.

### Methods details

#### Clinical Assessments and Data Collection

Disease severity was assessed with the ALSFRS-R. The progression rate prior to enrollment was estimated using the DeltaFRS, calculated by subtracting the ALSFRS-R score at baseline assessment from a maximum score of 48, and dividing by the number of months passing from symptom onset to baseline assessment. Any use of riluzole was recorded. Index date (follow-up “time zero”) was defined as the date of enrollment and survival time was measured from the index date to events for either mortality, tracheostomy or permanent assisted ventilation (PAV). The censoring date for survival analysis was documented in a case where an event was not observed before the study ended. Experimenters were blinded during the steps of the molecular analysis. Whole blood was collected by venipuncture in ethylenediaminetetraacetic acid (EDTA) tubes, and plasma was recovered by centrifugation at room temperature (RT) for 10 min between 1300g-1750g. Plasma was stored at < −70_°C prior to use Clinical information for patients is shown in Table 1.

#### Small RNA next-generation sequencing

The molecular experimenters were blinded to the identity of the samples. Total RNA was extracted from 500μl plasma using miRNeasy Micro Kit following the manufacturer’s protocol except adding 2.5ml of Qiazol to each plasma sample (Qiagen, #217084). Samples were homogenized by a 30 second vortex. No biological or technical repeats were used. RNA was quantified with a Qubit fluorometer using the RNA Broad Range Assay Kit (Thermo Fisher Scientific, #Q10211) following the manufacturer’s protocol. Small RNA next-generation sequencing libraries were prepared using the QIAseq miRNA Library Kit (Qiagen, #331502) from 7.5_ng of total RNA following the manufacturer’s protocol. If RNA concentration was below the limit of detection, 5μl of undiluted RNA were used.

Precise linear quantification of miRNAs was achieved by 12-nucleotide long UMIs^107^. Samples were randomly assigned to the preparation of libraries indexed with QIAseq miRNA NGS 48 Index IL (Qiagen, #331595) and sequenced in batches of 48 or fewer samples. Library concentration was determined with a Qubit fluorometer following the manufacturer’s protocol (dsDNA High Sensitivity Assay Kit, Thermo Fisher Scientific, #Q32854) and library size with TapeStation D1000 (Agilent). Libraries with different indices were multiplexed and sequenced on a NextSeq 500/550 v2 (Illumina, #20024906) or a NovaSeq 6000 flow cell (Illumina, #20028401), with a 75-bp single read or paired-end reads with 6-bp index. FASTQ files were de-multiplexed using the user-friendly transcriptome analysis pipeline^108^. Human miRNAs and isomiRs, as defined by miRBase - V22^109^, were mapped using CLC genomic workbench 22.0 (Qiagen) only to the positive strand. Reads were trimmed in two consecutive fashions; Phred score^110^, based error () was calculated for every nucleotide, high values reflect poor read quality. A trimming score defined by the subtraction of the error from a threshold of 0.05 was set for each nucleotide. Then, a running sum of the nucleotide errors was calculated per read. The region between the first positive value of the running sum and the highest value of the running sum was kept with non-positive score reads being removed completely. Consecutively, reads’ ends were trimmed to keep the maximum length region containing 2 or fewer ambiguous nucleotides. This was followed by the library’s 3’-adaptors (TGGAATTCTCGGGTGCCAAGG) trimming, with reads that did not contain any adaptors being excluded from downstream analyses. Reads were filtered based on lengths of 15-55 bps. IsomiRs were considered as 2bps changes (insertion, deletion, shift, and mismatch) up/downstream from miRBase reference sequence with a maximum of 2 mismatches allowed in alignment. The algorithm prioritizes perfect matches to reference miRNAs, followed by sequences of increasing mismatch nucleotides. When a sequence aligns equally well to multiple isomiRs, counts are assigned uniformly across all matching forms.

#### IsomiR definitions

Reference miRNA refers to the most abundantly expressed sequence variant among all isomiRs derived from a given miRNA gene. The collective set of all isomiRs, often comprising around 50 distinct variants annotated for a specific miRNA gene is defined as the miRNA.

#### QC and pre-processing of sequencing data

Sequencing data were pre-processed separately for each cohort (British n=154 / PGB n=200) and yielded ∼280K/∼228K isomiRs, respectively, but 99% of identified isomiRs were extremely rare and sparsely detected across samples. Therefore, isomiRs were included in the analysis only f their average expression level across cohort’s samples was above 5 UMIs, yielding a total of 2046 and 2003 isomiRs in the British and PGB cohorts, respectively. As geometric mean cannot be calculated with missing values, the remaining isomiR missing reads were imputed using multiple imputation with a denoising autoencoder algorithm (parameters: 40 forward passes, three layers with 512 nodes each, probability of corruption of 0.95, learning rate of 1e^-6^). Missing values were replaced by the mean isomiR levels across 50 imputed datasets. Clinical features used for estimation of similarity between patients: age, sex, site of symptom onset, and Riluzole treatment. Python-V3.9.2 and R-V4.0.4 package *rMIDAS*-V.0.3^111^ were used for the imputation. Using *DESeq2*^112^, libraries were corrected for size by geometric mean, under the assumption that miRNA counts followed negative binomial distribution^112^. Then, imputed values of original missing values were transformed back to zero to avoid bias in batch correction. Finally, data were corrected for the library preparation batch to reduce its potential bias by the R Combat function in *SVA*-V3.38 package^113^. Then, levels of 1480 isomiR identities shared across the two cohorts were standardized and isomiRs were considered for downstream analyses.

#### Machine learning-based isomiR selection for disease prognostication

Biomarker discovery analysis (training) was performed using the UK cohort that we have recently characterized^17^ while the PGB cohort was reserved for external validation. The 1,480 isomiR candidates were randomly sampled with repetitions to generate 3,718 bootstrap subsets, each containing 20 isomiRs. The sampling process ensured that each isomiR was included in at least 50 distinct subsets. Importantly, any given subset contained one specific isomiR of interest alongside 19 other randomly selected isomiRs, resulting in at least 50 different subset compositions for each individual isomiR. For each bootstrap set survival analysis was conducted using three survival models: Cox regression analysis^42^, RSF^44^, and XGB^43^. The analyses were performed using the scikit-survival library (version 0.23.0) in Python. SFS^114^, adds iteratively isomiRs in different combinations to optimize the C-index^115^ (*mlxtend* library in Python).

The process explored approximately 1,000,000 possible combinations of isomiRs for each subset , resulting in roughly 3.9 billion combinations across all 3,718 isomiRs subsets. Each isomiR combination was evaluated using 10-fold CV on subsets of the UK discovery cohort within each of the three orthogonal survival models. A selection threshold of ≥90% in the SFS process yielded 84 isomiRs. Subsequently, additional iterations of feature selection were performed on the 84 isomiRs, generating 222 random subsets for further analysis. A C-index-based threshold was determined using the elbow method^53^ and applied across all random subsets. This process identified two final panels, together consisting of 18 unique isomiRs, which were selected for downstream analyses.

#### isomiRs panel prediction validation

After feature selection, an XGB model was trained on the UK discovery cohort to predict patient survival time from enrollment. Hyperparameter tuning was conducted using 5-fold CV over three rounds of *Optuna* optimization^116^ (1000 trials per round, with a random sampler and seed = 42), followed by manual fine-tuning. The optimized model was trained on the selected isomiR levels from the UK cohort and validated using the corresponding isomiR levels from the PGB replication cohort. SHAP^117^ were used to identify feature importance within the model and were calculated on the individual predicted hazard function. HRs, based on SHAP values, were calculated as previously described^54^. Briefly, Individual SHAP values were exponentiated across features and values were averaged separately for all patients below feature mean level and those above it. The average value of above mean levels was divided by the average value of below mean level to yield the hazard ratio. The performance of the chosen isomiR-based model was compared against alternative models trained on: reference miRNA levels only, miRNAs levels (defined as the aggregate of all isomiRs within a miRNA annotation).

#### NfL assay

NfL levels were quantified using single molecule array (Simoa) technology on the Simoa HD-1 Analyzer (Quanterix) in plasma/serum samples from 152 and 200 patients in the UK discovery and PGB replication cohorts, respectively. The assay utilized the Simoa NfL Advantage Kit (Quanterix, #102258), with standards, primary and secondary antibodies, and detection ranges specified by the manufacturer, including defined lower and upper detection limits. Univariate or multivariate Cox regression, with or without clinical features, and associations between these and mortality were analyzed as described below.

#### Feature engineering

For univariate/multivariate Cox regressions isomiR levels and NfL levels were dichotomized into binary variables based on the median levels in the UK discovery cohort. miR-181 levels were dichotomized using a previously published threshold of 71,000 UMIs ^17^. Combinations of isomiR-let-7g-5p.t with NfL or miR-181 were evaluated by their ratios, followed by a dichotomization based on the median levels in the UK discovery cohort for NfL combination, or by an in-house algorithm that evaluated dichotomization cutpoint across all possible values by optimizing univariate Cox mortality HR on the UK discovery cohort.

#### miRNA target prediction

mRNA targets of the reference miRNA were retrieved from miRCarta^55^, retaining those listed in at least one of miRTarBase, micro-T-CDS, or TargetScan. Target canonical transcripts sequences were obtained from Ensembl^118^ using *BioPython* V-1.83^119^. miRNA and mRNA sequences were analyzed using RNA22-v2^56^ with a user defined sensitivity-optimized parameters (see Supplementary Methods). Briefly, lower values of minimum base-pair matches and higher values of free energy were set to increase prediction of non-canonical targets in addition to allowing 2 mismatches at the miRNA-mRNA seed pairing and G:U wobbling. Moreover, a higher target detection threshold was chosen based on predefined program setting. Significant hybridization sites were counted and compared between the reference miRNA and the isomiR. Targets with differing site counts were analyzed for enrichment using Enrichr^120–122^ via *GSEApy V-1.1.9*^123^ and tested against an ALS gene list obtained from Eitan et al^124^.

#### Quantification and Statistical Analysis

The Student’s t-test was applied to assess significant differences in the age at symptom onset or study enrollment between groups. The Mann–Whitney U test was used to evaluate differences in ALSFRS-R, DeltaFRS scores, and isomiR-let-7g-5p.t levels between the groups. Differences in the proportions of censored events, sex prevalence, or Riluzole treatment were tested using a chi-squared goodness-of-fit test. One-way ANOVA with Tukey-HSD post-hoc analysis assesses differences in isomiR-let-7g-5p.t levels across ALS sub cohorts and control. The Shapiro-wilk test was used to determine normal distribution. The Wilcoxon signed-rank test was used to evaluate enrichment of miRNA targets for ALS genes. Pearson correlation coefficients were calculated to examine the relationships between isomiR levels within/between the two cohorts. Kaplan–Meier survival estimators were analyzed using the log-rank Mantel–Cox test, with survival data censored at a fixed date. Univariate and multivariate Cox proportional hazard models were employed to estimate mortality hazard ratios from the time of enrollment, and model fit was assessed using the C-index. Proportional hazard assumptions were assessed by R package *survival* function using the cox.zph function. The performance of the XGB model was evaluated using the dynamic AUC(t)^125^ across the range of survival periods, the average AUC(t), and the C-index. Specific statistical tests, relevant N values and their definition, definition of center, and dispersion measures were reported in Tables, and Figure and Table legends. No sample size estimations were made in this study. All statistical analyses were conducted, and visualizations were generated using Python (version 3.9.2), R (version 4.0.4), and BioRender.

### Parameters to fit models for feature selection

Models for were fitted with the following:

- Cox: without regularization, ties = ’breslow’ and a tolerance of 1e-9.
- Random Survival Forest: n_estimators = 2, max_depth = 4, max_samples = 0.25, n_estimators = 20, min_samples_split = 2, min_samples_leaf = 2, max_features = 0.5, max_samples = 0.5, min_weight_fraction_leaf = 0.0, max_leaf_nodes = None, bootstrap = True, random_state = 42).
- Survival Gradient Boosting: n_estimators = 20, learning_rate = 0.5, max_depth = 2, subsample = 0.3, loss = ’coxph’, criterion = ’friedman_mse’, min_samples_split = 2, min_samples_leaf = 1, min_weight_fraction_leaf = 0.0, min_impurity_decrease = 0.0, max_features = None, max_leaf_nodes = None, validation_fraction = 0.1, n_iter_no_change = None, tol = 0.0001, dropout_rate = 0.3, ccp_alpha = 0.0,random_state = 0)

### Parameters to fit Tuned survival gradient boosting hyperparameters

n_estimators = 95, learning_rate = 0.1, max_depth = 35 subsample = 0.4545454545454545, loss = ’coxph’, criterion = ‘friedman_mse’, min_samples_split = 60, min_samples_leaf = 20, min_weight_fraction_leaf = 0.060461799614030046, min_impurity_decrease = 0.6, max_features = 0.1111111111111111, max_leaf_nodes = 100, validation_fraction = 0.1, n_iter_no_change = None, tol = 0.0001, dropout_rate = 0.016, verbose = 1, ccp_alpha = 0.1, random_state = 42.

### Parameters for increased sensitivity analysis of RNA22 v2

Seedmismatchsallowed = 2, seedsize = 8, minpairmatchs = 10, minenergy = -10, maxguwobbles = - 1, and threshold = 3000000

