## Supplementary figures + legend for "IsomiR Utility in Amyotrophic Lateral Sclerosis Prognostication"

**Table S1**

| Mutation | Nomenclature | Reference sequence | Mutated sequence | Comment |
|---|---|---|---|---|
| **Reference** | Let-7g-5p | TGAGGTAGTAGTTTGTACAGTT | TGAGGTAGTAGTTTGTACAGTT | No change |
| **3' deletion** | Let-7g-5p.tt | TGAGGTAGTAGTTTGTACAG**TT** | TGAGGTAGTAGTTTGTACAG | Lowercase letter (n) |
| **3' insertion** | Let-7g-5p.CC | TGAGGTAGTAGTTTGTACAG | TGAGGTAGTAGTTTGTACAG**CC** | Uppercase letter (N) |
| **5' deletion** | Let-7g-5p.stg | **TG**AGGTAGTAGTTTGTACAGTT | AGGTAGTAGTTTGTACAGTT | Addition of "s" |
| **5' insertion** | Let-7g-5p.sGG | TGAGGTAGTAGTTTGTACAGTT | **GG**TGAGGTAGTAGTTTGTACA GTT | Addition of "s" |
| **Base exchange** | Let-7g-5p.G2C.T16G | TGAGGTAGTAGTTTGTACAGTT | T**C**AGGTAGTAGTTTG**G**ACAGTT | Position relative to reference sequence |

**Table S1 - IsomiR Nomenclature differs from miRNA**, related to figure 2A. Mutations are separated by a dot. Examples of mutations are highlighted in bold and red.
Base exchanges are represented by the original nucleotide, followed by its position in the reference sequence, and then the new nucleotide (e.g., A12G).
Reference miRNAs do not have any suffixes. Deletions are indicated by lowercase letter suffixes. Insertions are indicated by uppercase letter suffixes.
5′ indels are prefixed with the letter "s"; otherwise, indels are assumed to be at the 3′ end. The position of base exchange in indels remains relative to the reference sequence.

| Feature | C-index | HR | 95% CI Lower | 95% CI Upper | P | C-index | HR | 95% CI Lower | 95% CI Upper | P |
|---|---|---|---|---|---|---|---|---|---|---|
| cohort | UK - discovery | | | | | PGB - replication | | | | |
| miRNA-let-7g-5p | 0.51 | 0.88 | 0.63 | 1.22 | 0.43 | 0.51 | 1.08 | 065 | 1.82 | 0.76 |
| miRNA-let-7a-5p | 0.51 | 1.39 | 0.99 | 1.94 | 0.06 | 0.59 | 0.73 | 0.46 | 1.18 | 0.2 |
| miRNA-191-5p | 0.52 | 0.76 | 0.55 | 1.06 | 0.11 | 0.50 | 4.0 | 0.4 | 12 | 0.99 |
| miRNA-223-3p | 0.53 | 0.71 | 0.51 | 0.99 | 0.05 | 1.0 | 1.0 | 1.0 | 1.0 | 1.0 |
| miRNA-144-3p | 0.53 | 0.79 | 0.57 | 1.10 | 0.16 | 0.53 | 1.34 | 0.88 | 2.05 | 0.17 |
| miRNA-185-5p | 0.53 | 0.69 | 0.49 | 0.96 | 0.03 | 0.5 | 1.0 | 1.0 | 1.0 | 1.0 |
| miRNA-339-5p | 0.53 | 0.83 | 0.6 | 1.15 | 0.25 | 0.5 | 1.0 | 1.0 | 1.0 | 1.0 |
| miRNA-141-3p | 0.57 | 0.65 | 0.47 | 0.91 | 0.01 | 0.5 | 1.11 | 0.71 | 1.73 | 0.66 |
| miRNA-16-5p | 0.51 | 0.92 | 0.66 | 1.27 | 0.61 | 0.51 | 1.78 | 0.24 | 12.9 | 0.57 |
| miRNA-221-3p | 0.52 | 0.83 | 0.6 | 1.16 | 0.27 | 0.5 | 0.77 | 0.11 | 5.59 | 0.8 |
| miRNA-20a-5p | 0.59 | 0.49 | 0.35 | 0.68 | 3.2e-05 | 0.5 | 1.0 | 1.0 | 1.0 | 1.0 |
| miRNA-363-3p | 0.52 | 0.89 | 0.64 | 1.23 | 0.48 | 0.56 | 1.74 | 1.13 | 2.66 | 0.01 |
| miRNA-19a-3p | 0.58 | 0.46 | 0.32 | 0.65 | 1.2e-05 | 0.5 | 4.0 | 0.4 | 12.0 | 1.0 |
| miRNA-30e-5p | 0.55 | 0.57 | 0.4 | 0.8 | 1.1e-3 | 0.5 | 1.4 | 0.19 | 10.1 | 0.74 |

**Table S2 – miRNAs, a term reflecting on the integration of all isomiRs associated to a miRNA gene, display poor prognostication capacity**, related to Figure 2C-D. Cox univariate results of the miRNA associated to isomiRs from Figure 2. Results are shown for UK – discovery cohort (blue) and PGB – replication (purple). P – p-value, HR – hazard ratio, C-index – concordance index, CI – confidence interval.

| Name | miRNA UK discovery | miRNA PGB replication | reference miRNA UK discovery | reference miRNA PGB replication |
|---|---|---|---|---|
| **hsa-let-7e-5p** | 545.76 ± 350.51 | 561.41 ± 244.80 | 290.93 ± 225.13 | 303.17 ± 137.28 |
| **hsa-let-7f-5p** | 9434.27 ± 4849.83 | 11554.54 ± 4143.22 | 5908.90 ± 4210.59 | 7920.46 ± 3218.58 |
| **hsa-let-7g-5p** | 992.89 ± 372.96 | 1173.12 ± 307.08 | 555.03 ± 385.22 | 713.87 ± 264.32 |
| **hsa-let-7i-5p** | 12677.13 ± 5929.79 | 14073.11 ± 4257.76 | 9116.78 ± 5659.07 | 9864.22 ± 3331.09 |
| **hsa-miR-130b-5p** | 8.26 ± 5.39 | 9.40 ± 4.97 | nan | nan |
| **hsa-miR-142-3p** | 1180.20 ± 776.49 | 2430.42 ± 1049.35 | 61.75 ± 44.68 | 123.52 ± 63.52 |
| **hsa-miR-144-3p** | 24.58 ± 27.28 | 16.31 ± 18.94 | 8.63 ± 9.66 | 5.56 ± 6.45 |
| **hsa-miR-146a-5p** | 2270.01 ± 1118.95 | 6507.35 ± 1719.87 | 1200.03 ± 835.11 | 3592.34 ± 1241.20 |
| **hsa-miR-151a-3p** | 1480.06 ± 677.72 | 4782.99 ± 1337.09 | 802.72 ± 467.30 | 2963.98 ± 869.57 |
| **hsa-miR-25-3p** | 1865.72 ± 1072.00 | 2025.82 ± 669.98 | 1363.26 ± 744.40 | 1630.85 ± 599.57 |
| **hsa-miR-30d-5p** | 2920.10 ± 861.54 | 4553.77 ± 999.95 | 275.84 ± 146.96 | 399.31 ± 133.34 |
| **hsa-miR-425-5p** | 865.75 ± 427.35 | 914.48 ± 244.07 | 129.94 ± 90.52 | 213.61 ± 80.55 |
| **hsa-miR-432-5p** | 653.12 ± 706.14 | 640.87 ± 407.83 | 241.62 ± 214.92 | 266.80 ± 176.47 |
| **hsa-miR-451a** | 1416.05 ± 1993.90 | 550.35 ± 745.91 | 257.16 ± 461.63 | 133.97 ± 225.36 |
| **hsa-miR-486-5p** | 25375.59 ± 29731.65 | 7693.64 ± 7295.35 | 9280.17 ± 8542.15 | 2768.21 ± 2999.47 |
| **hsa-miR-92a-3p** | 7033.59 ± 6283.45 | 4454.01 ± 2302.21 | 4381.68 ± 3579.75 | 3375.72 ± 1824.18 |

**Table S3 – plasma levels of reference miRNAs and miRNAs (integrated of all isomiRs associated to a defined miRNA gene) associated to the 18 isomiRs found by ML approach**, related to Figure 3B-E. Mean plasma levels and standard deviation, in UMIs, displayed across patients in the UK discovery and the PGB replication cohorts..

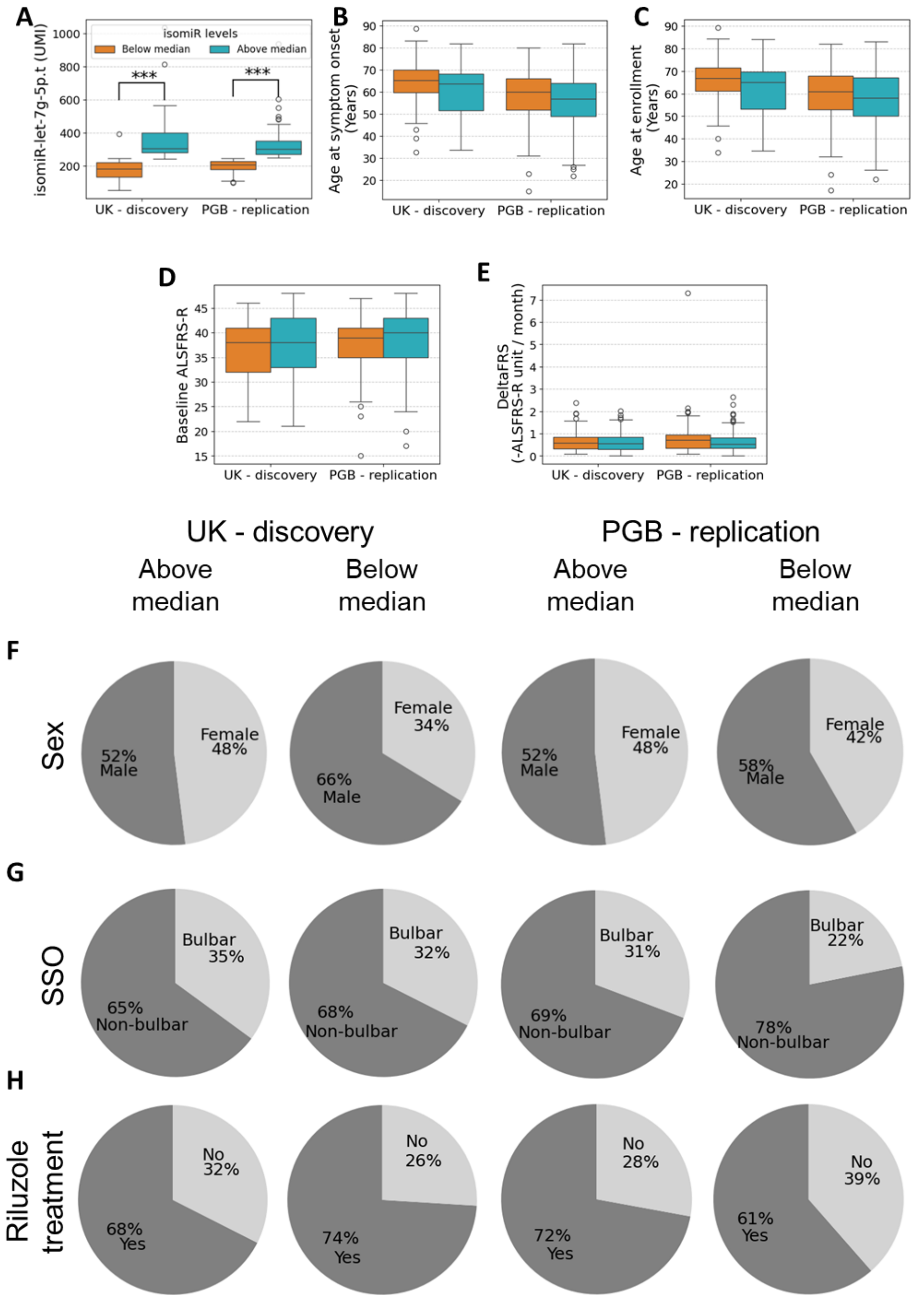


**Figure S1 – Above/below median levels of isomiR-let-7g-5p.t are not correlated to clinical features**, related to Figures 1 and 2E-F. Boxplots of **(A)** isomiR-let-7g-5p.t levels, **(B)** Age at disease symptom onset, **(C)** Age at study enrollment**,** **(D)** baseline ALSFRS-R score or **(E)** DeltaFRS, measured as the reduction in the ALSFRS-R score per month in above/below median groups across cohorts. Pie charts of **(F)** Sex, **(G)** SSO - site of symptom onset or **(H)** Riluzole treatment above/below median groups across cohorts. *** P-value ≤ 0.001, two-sided Mann-Whitney U test. The $1^{st}$ and $99^{th}$ percentiles of isomiR levels are denoted by whiskers. Extreme values are marked by circles. $N_{UK\ above}$ = 77, $N_{UK\ below}$ = 77, $N_{PGB\ above}$ = 160, $N_{PGB\ below}$ = 40.

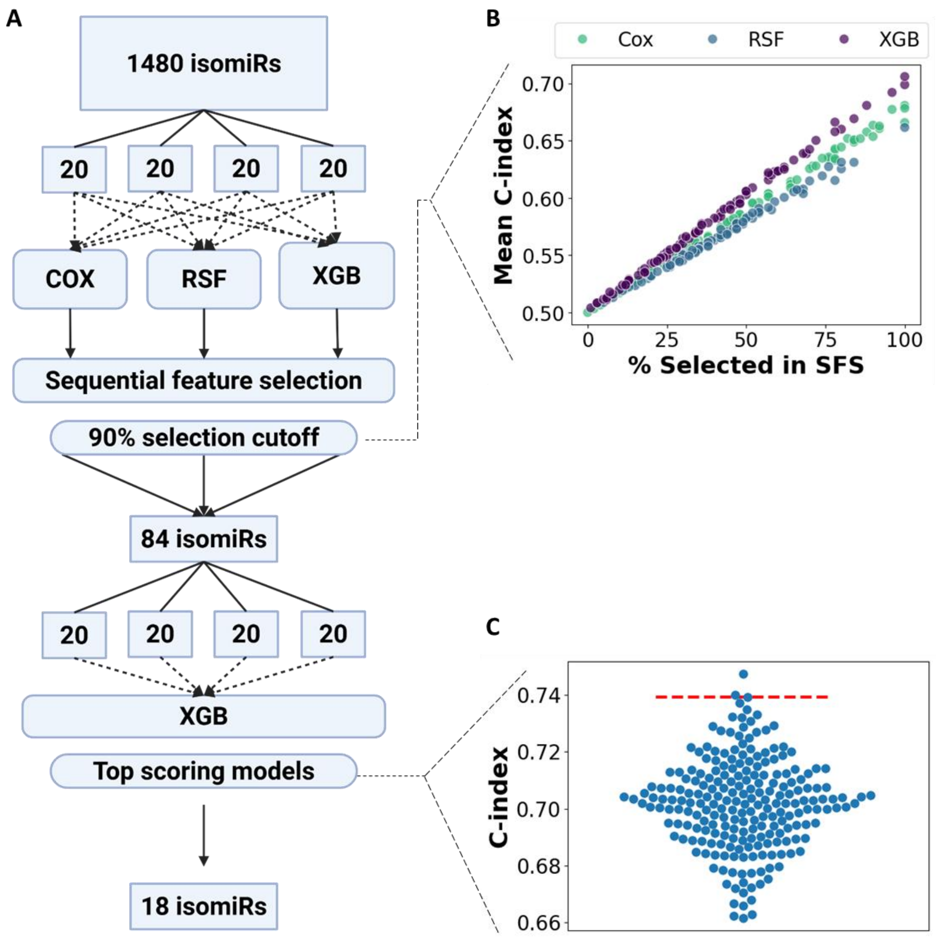


**Figure S2 - Feature selection process yielded 18 candidate isomiRs as optimal predictors for survival**, related to Figure 3A. **(A)** Schematic overview of the multi-step feature selection strategy applied to 1,480 isomiRs profiled in a UK discovery cohort (N = 154). Initially, three different survival models, Cox proportional hazards (COX), Random Survival Forest (RSF), and gradient survival boosting (XGB), were each applied across repeated data splits, composed of 20 isomiRs. For each model, sequential feature selection (SFS) was performed to identify isomiRs contributing most to predictive performance. IsomiRs selected in at least 90% of the SFS runs were retained, yielding a set of 84 highly stable candidates. This refined set underwent a second round of XGB modeling. The top-scoring models from this phase were based on concordance index (C-index). **(B)** Scatter plot of the mean C-index (n = 50) across 3,718 isomiR subsets (y axis) and percentage of times isomiRs were selected in sequential forward selection (SFS) in three survival analyses: Cox (green), RSF (blue), or XGB (purple). **(C)** A plot of C-index (y axis) for sampling of 20 isomiRs from a total of 84, with repetition, using XGB (n = 222). Top two models, above the red dashed line, displayed the best C-index.

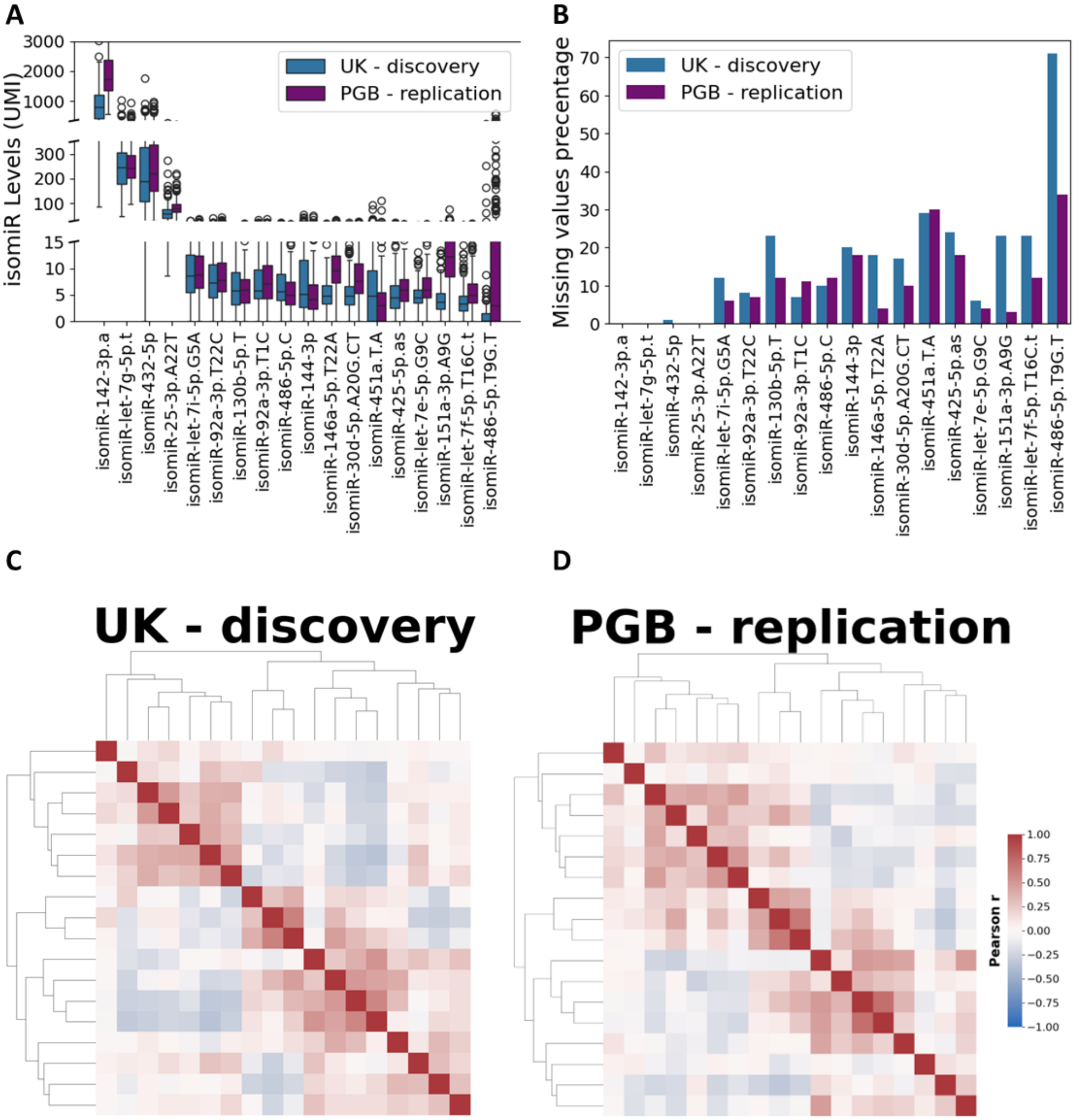


**Figure S3 – isomiR characteristics are similar across the discovery and replication cohorts**, related to Figure 3F-H **(A)** Boxplot of corrected isomiR levels (y-axis) in the UK discovery cohort (blue, N = 154) and the PGB replication cohort (purple, N = 200). The 1$^{st}$ and 99$^{th}$ percentiles of isomiR levels are denoted by whiskers. Extreme values are marked by circles **(B)** Bar plot of missing isomiR values in patients from the UK discovery (blue) and PGB replication (purple) cohorts. Heatmaps of the correlation between plasma isomiR levels in **(C)** the UK discovery cohort and **(D)** the PGB replication cohort clustered by hierarchical clustering according to Euclidean distance and average linkage by UK discovery data. Pearson's correlation r from -1.0 (complete negative correlation, blue) to 1.0 (complete positive correlation, red).

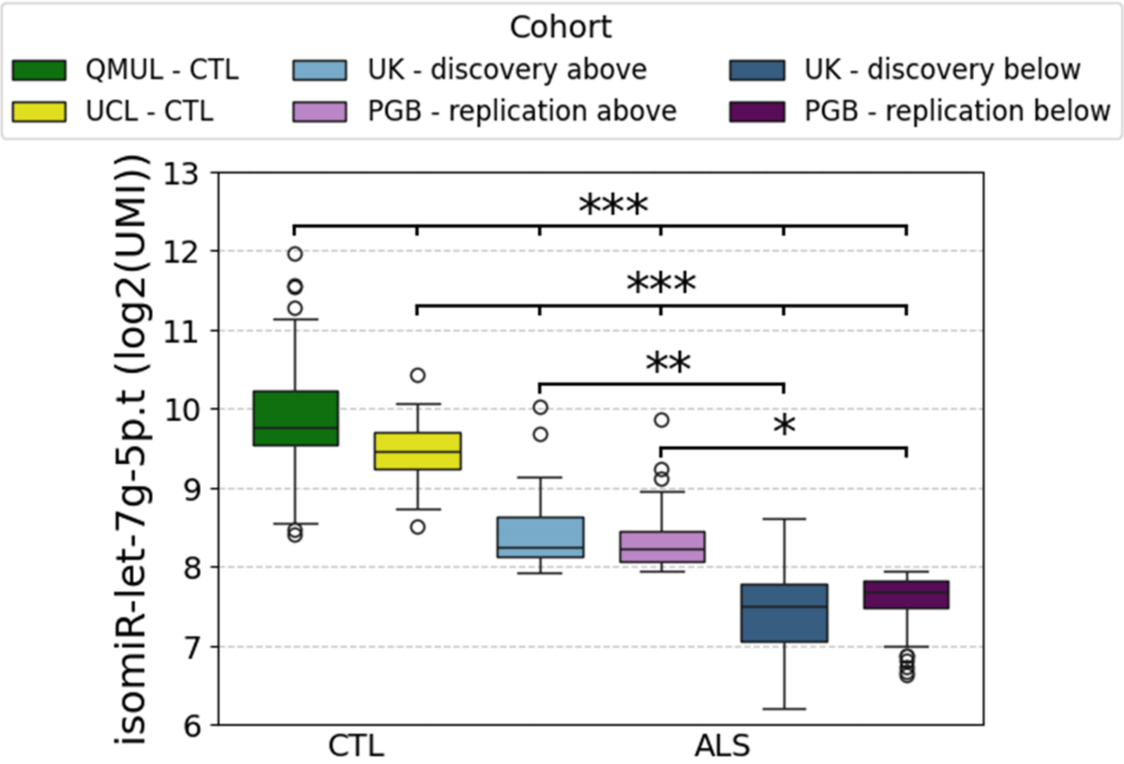


**Figure S4 – isomiR-let-7g-5p.t plasma levels are reduced in ALS patients compared to controls**, related to Figure 2C-F. Boxplot of isomiR-let-7g-5p.t levels in plasma of controls from two independent cohorts (green , N = 99; yellow, N = 56) and ALS sub cohorts above, (light blue, n = 77; light purple, N = 160) or below (dark blue, N = 77; dark purple, N = 40) in the UK discovery or PGB - replication cohorts, respectively. * P-value ≤ 0.05, ** P-value ≤ 0.01 , *** P-value ≤ 0.001, Tukey's HSD post-hoc test. The 1st and 99th percentiles of isomiR levels are denoted by whiskers. Extreme values are marked by circles.

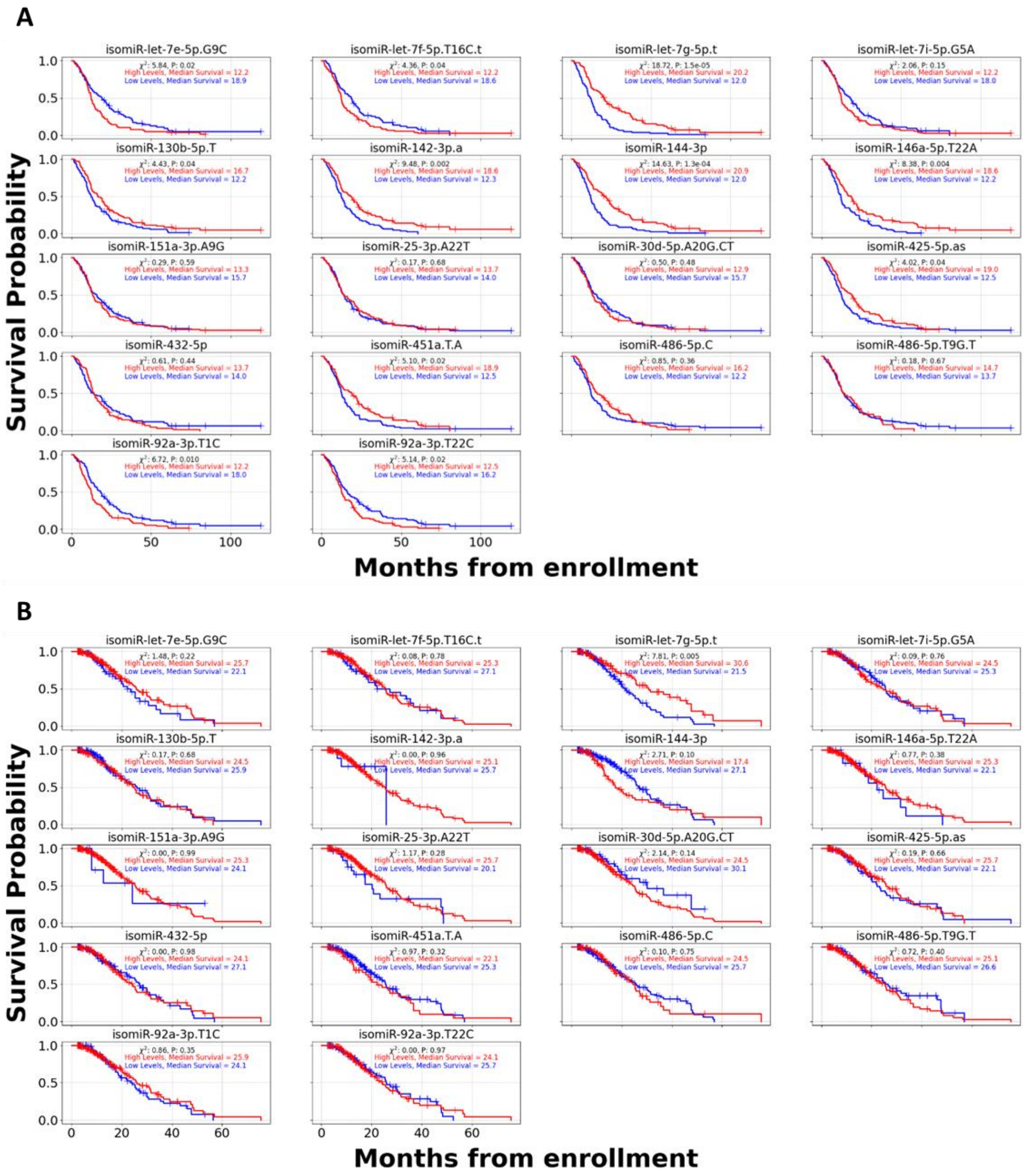


**Figure S5 - Cumulative survival (Kaplan–Meier) curves for ML selected isomiRs illustrate the prognostic importance of isomiR-let-7g-5p.t**, related to Figure 3F-H. Data dichotomized by UK discovery isomiR median values. Curves represent values from the **(A)** UK - discovery or **(B)** PGB – replication cohorts, respectively. Subthreshold or suprathreshold groups are denoted in blue / red, respectively. Group median survival and Logrank statistics are presented.

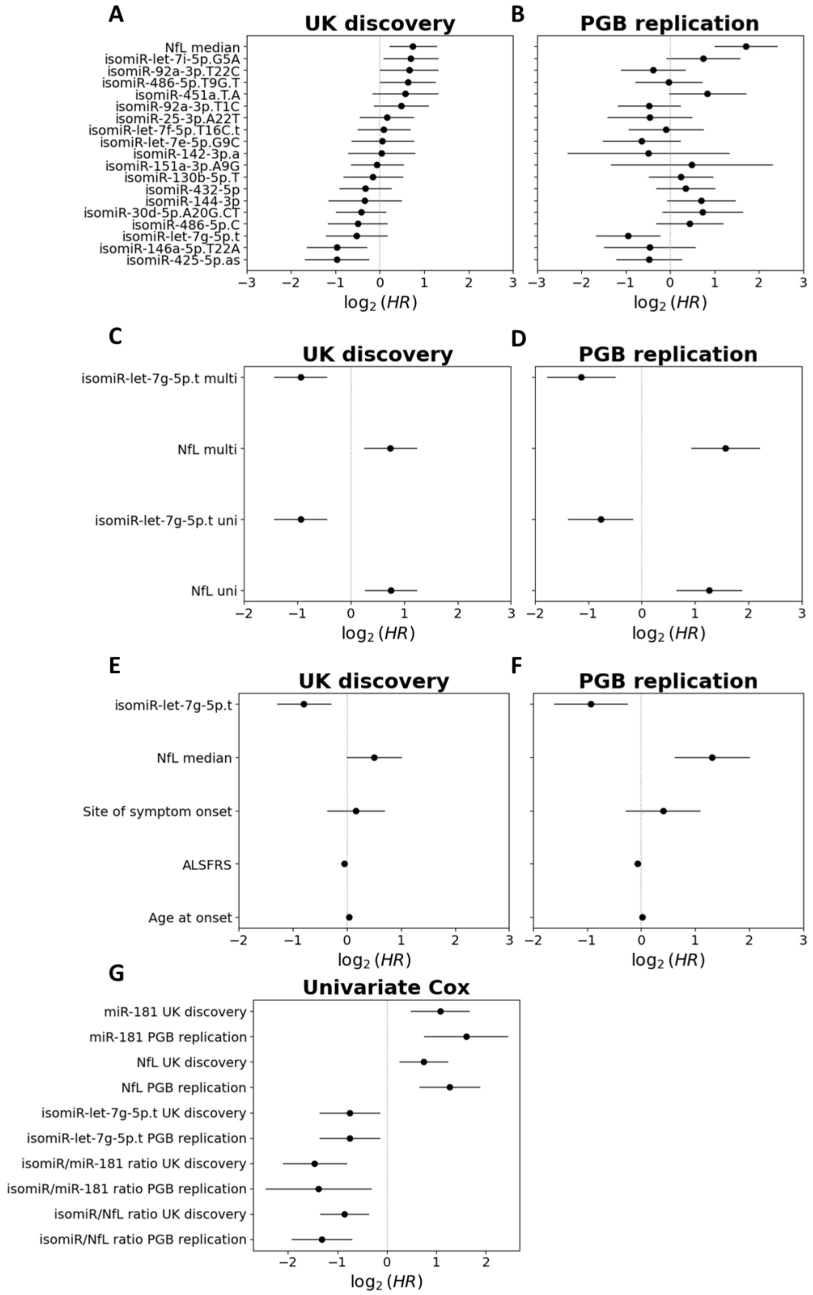


**Figure S6 – Improvement of prognostic performance of joint isomiR-let-7g-5p.t and NfL signal**, reacted to Figure 4. Forest plots of multivariate Cox proportional hazard ratios of 18 isomiRs and NfL in **(A)** the UK discovery or **(B)** PGB replication cohorts. Multivariate and univariate Cox hazard ratios of NfL and isomiR-let-7g-5p.t in the UK discovery **(C)** or PGB replication **(D)** cohorts Multivariate Cox hazard ratios of NfL, isomiR-let-7g-5p.t and clinical features in the UK discovery **(E)** or PGB replication **(F)**. Univariate Cox proportional hazard ratios of isomiR-let-7g-5p.t, NfL, miR-181 and the ratios of isomiR-let-7g-5p.t,/NfL or isomiR-let-7g-5p.t/miR-181, in both cohorts **(G)**. HR CI denoted by horizontal line.
